# Enhancer-targeting CRISPR screens at coronary artery disease loci suggest shared mechanisms of disease risk

**DOI:** 10.1101/2025.08.28.25334684

**Authors:** Markus Ramste, Chad Weldy, Soumya Kundu, Quanyi Zhao, Daniel Li, Kayla Brand, Disha Sharma, Amanda Ramste, Evelyn Jagoda, Judhajeet Ray, Roxanne Diaz Caceres, James Galante, Andreas R. Gschwind, Nuutti Lahtinen, Trieu Nguyen, Junedh M. Amrute, Chong Yong Park, Juyong Brian Kim, Minna U Kaikkonen, Nathan O. Stitziel, Lars Steinmetz, Anshul Kundaje, Jesse M. Engreitz, Thomas Quertermous

## Abstract

To systematically identify causal genetic mechanisms that confer risk for coronary artery disease (CAD) in GWAS loci, we mapped genome-wide variant-to-enhancer-to-gene (V2E2G) links in vascular smooth muscle cells (SMC). Enhancers identified by active chromatin features, and further prioritized by base-resolution deep learning models of chromatin accessibility in 108 CAD loci, were studied with CRISPRi targeting and Direct-Capture Targeted Perturb-seq (DC-TAP-seq) evaluation of 470 genes. Seventy-six V2E2G links were identified for 59 candidate CAD genes representing gene programs including epithelial-mesenchymal transformation, ubiquitination, and protein folding as well as BMP and TGFB signaling. Similar methods employed with an independent focused screen targeting one candidate locus at 9p21.3 identified 10 enhancers regulating expression of multiple genes at this location. Detailed molecular studies revealed that two enhancers mediating transcription factor binding and transcriptional regulation contribute to ancestry-specific and sex-specific risk for CAD and the surrogate biomarker vascular calcification. Together, these studies advance our identification of GWAS CAD V2E2G links across the genome, and specific mechanisms of risk at the complex 9p21.3 locus.

## Introduction

Coronary artery disease (CAD) is the worldwide leading cause of human mortality ^1^. As with other complex diseases, CAD is due to aberrant changes in gene expression, due to complex regulatory epigenetic and transcriptomic mediated cellular responses to vascular stresses. With the identification of over 300 replicated loci, genome wide association studies (GWAS) have provided the first opportunity for the identification of cellular and molecular pathways that regulate genetic risk for CAD ^2,3^. Much of this risk resides in the coronary smooth muscle cell (SMC) lineage, as determined by algorithms incorporating recent CAD meta-analysis association data and relevant tissue gene expression or chromatin accessibility data ^3–7^. Studies in this laboratory have thus focused on SMC, and have successfully identified and linked a small number of causal genes to complex phenotypic transitions in this cell type. We have shown that genes protective toward disease risk, such as *TCF21* and *SMAD3,* promote the transition of SMCs to a fibroblast-like fibromyocyte (FMC) phenotype and inhibit transition to a chondrogenic chondromyocyte (CMC) phenotype, while risk promoting genes like *PDGFD* contribute to the development of CMC ^8–11^. Work in this laboratory and others investigating risk at a genome-wide level have suggested that the CAD risk is encoded in genes that modulate these SMC state transitions ^12^. Further studies linking specific causal genes will be required to investigate in a comprehensive fashion how these genes modulate SMC phenotypic transitions that are associated with disease related molecular mechanisms. Also, study of the epigenetic basis of variation in causal CAD gene expression can provide insights into mechanisms by which racial-ethnic and sex differences in disease risk are encoded in specific loci.

The challenge of using GWAS data to investigate cellular and molecular mechanisms of complex disease risk resides in the difficulty to identify causal genes within the associated loci. Fine mapping and prioritization approaches using various types of genomic data have not provided reliable identification of causal genes. However, recent use of CRISPR methodology to perturb enhancer function promises to provide a significant advance in the field, allowing high throughput identification of genes whose expression is regulated by a broad range of enhancers across the genome ^13–15^. In this approach, multiple guide RNAs (gRNAs) are introduced into relevant cultured cells, and activation of dCas9-CRISPRi constructs leads to the knockdown of enhancer regulated genes, which are then assessed through single cell RNA sequencing (scRNAseq). An important recent modification of this method incorporates polymerase chain reaction amplification and quantitation of disease locus genes, in conjunction with gRNA detection (Direct-Capture Targeted Perturb-seq (DC-TAP-seq)) ^16^. This approach provides significant improvement in sensitivity of the methodology. While enhancer-Perturb-seq in general, and the DC-TAP-seq method specifically, have shown great promise in experimental model systems, this approach has not been widely used to create variant to enhancer to gene (V2E2G) links for complex diseases such as CAD.

In studies reported here we have employed immortalized human coronary artery cells (HCASMC) expressing dCas9-KRAB-ZIM3 to screen 209 enhancers in 108 CAD loci to identify 69 CAD genes. These genes identified cellular programs including osteochondrogenic development, epithelial-mesenchymal transformation, ubiquitination, protein folding, and BMP and TGFB signaling. A focused CRISPRi screen of the 9p21.3 locus identified 10 enhancers that were shown to regulate expression of multiple genes in this locus that regulate SMC phenotype. Further, detailed study of causal variation in two of these enhancers revealed transcriptional mechanisms that likely contribute to ancestry- and sex-related differences in CAD risk at this region of the genome. Overall, these studies address a critical need in the field to expand the number of causal genes in GWAS associated loci and link them together in specific programs, thus extending available knowledge and promoting further mechanistic studies to validate and explore their relevant biology.

## Results

### Characterization of human CAD associated enhancers that contain prioritized functional disease variation (V2E)

To allow Perturb-seq screens, we generated human telomerase reverse transcriptase (hTERT) immortalized human coronary artery smooth muscle cells (telo-HCASMC) which were transduced with lentivirus containing a dCas9-KRAB-ZIM3-P2A-mCherry construct for CRISPR interference (CRISPRi). These cells were FACS sorted and expanded to create a targeting HCASMC-CRISPRi cell line that was validated with gene-specific gRNAs.

To identify all variants associated with CAD, we obtained the genome-wide significant variants from multiple recent GWAS ^2,3,17^ and expanded that set to all common variants in high LD (r^2^ >0.8) using the 1000 Genomes European cohort. Next, to prioritize those variants that reside in regions with a likely functional role, we filtered the CAD-associated variants to those overlapping enhancers identified using pseudo-bulk scATAC-seq performed in the telo-HCASMC cell line. To further prioritize the enhancers that harbor variants that may have an allelic effect on chromatin accessibility, we trained ChromBPNet ^18^ models on mouse and human ^5^ in-vivo pseudo-bulk scATAC-seq from vascular tissues, along with pseudo-bulk scATAC-seq data from the telo-HCASMC cell line. Using those models, we evaluated the predicted effect of each CAD-associated variant on chromatin accessibility in both SMC and de-differentiated SMC cell states and retained only those variants predicted to induce a significant change in accessibility in either cell state. Finally, to ensure that the selected enhancers exhibited the potential to regulate nearby genes, we filtered for enhancers that met at least one of these criteria: located within 100KB of an expressed gene, within 250KB of a curated expressed gene, or linked to an expressed gene via Hi-C interaction ^19^ (**Fig. 1A)**.

**Figure 1.**
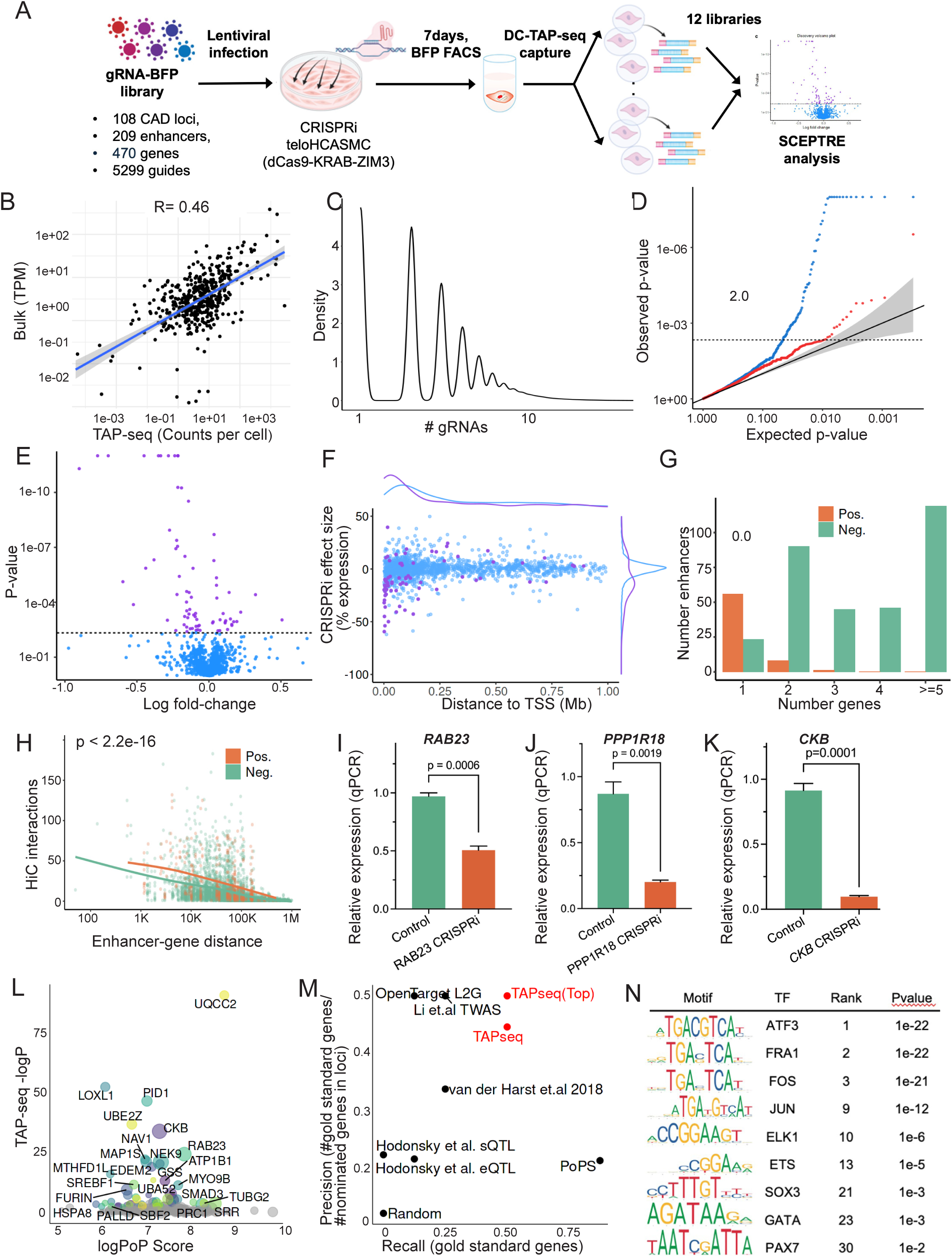
CRISPRi screen for genome-wide mapping of CAD GWAS enhancers to candidate causal genes. **A)** Design of SMC in vitro TAP-seq targeting screen of CAD enhancers. **B)** Correlation of gene expression assessed through the TAP-seq protocol in CRISPRi expressing telo-HCASMC compared to bulk RNA sequencing of native telo-HCASMC as assessed with bulk RNA sequencing. **C)** Graphical representation of guide RNAs (gRNAs, protospacers) transduced into CRISPRi-HCASMC (dCAS9-KRAB-ZIM3) by lentiviral infection. **D)** QQ plot comparing the distribution of experimental p-values compared to those expected to occur by chance. **E)** Volcano plot showing the log fold-change resulting from gRNA induced knockdown, represented as average fold-change per enhancer-gene pair. **F)** Representation of the distribution of causal gene effects in relation to enhancer distance from the transcription start site, with significant findings shown with red dots, and non-significant genes shown by grey dots, in relation to CRISPRi-HCASMC effect size as represented as % expression change. These V2E2G links have been uploaded to the Washington University Epigenome browser for visualization (https://shorturl.at/8P2qf). **G)** Number of genes linked to each CAD GWAS enhancer by DC-TAP-seq, and direction of effect for positive and negative effect sizes. **H)** Comparison of significant and non-significant E2G links for co-localization with HiC interactions (p<2.2e-16). **I, J, K)** CRISPRi-qPCR validation of TAP-seq V2E2G assignments for *RAB23, PPP1R18*, and *CKB*. **L)** Analysis with the polygenic priority score (PoPS) algorithm which learns trait-relevant gene features, such as cell-type-specific expression, to prioritize genes at GWAS loci ^28^. LogPoP score is compared with TAP-seq log P-value. **M)** Precision recall analysis to assess the ability of our DC-TAP-seq approach to identify gold standard SMC genes. **N)** Transcription factor motifs enriched at the V2E2G significant enhancers.

Prioritized variants were found to reside in 209 distinct candidate CAD enhancers in 108 unique genomic loci **(Fig. 1, Suppl. Table 1)**. For each of these enhancers, 8-15 targeting gRNAs were designed for each of the CAD loci **(Suppl. Table 1).** Primers were designed with CRISPRDesigner ^20,21^, to tile guides around that region, primarily targeting the summit of the ATAC peak. In addition, for each targeted CAD locus, we included both positive and negative control regions (Methods).

### In vitro SMC enhancer DC-TAP-seq multi-locus screens identify 59 unique variant to enhancer to gene (V2E2G) links

All genes with a transcription start site (TSS) within 100 kilobase pairs (kb) of the enhancers, linked to an enhancer via Hi-C data, or located within 250kb of at least one curated gene expressed in HCASMC with a TPM >10, were considered putative causal CAD genes for evaluation in the enhancer Perturb-seq studies (Suppl. Table 2). TAP-seq PCR amplification primers were designed using the previously reported customized pipeline ^16^ (**Suppl. Tables 3-4).** A total 5299 guides, including positive and negative controls, were cloned into lentivirus and transduced into the CRISPRi-HCASMC line, cells collected after 5 days with FACS using BFP fluorescence encoded by the guide containing lentivirus **(Suppl. Fig. 1A)**, and evaluated with ddPCR which showed a multiplicity of infection averaging ∼3 guides per cell. A total 270,790 isolated cells were divided among 12 individual 10x captures, libraries constructed and subjected to next-generation sequencing at an average depth of 17,256 reads per cell, and sequences mapped (**Suppl. Fig. 1B, C**).

The DC-TAP-seq approach provided good capture efficiency and statistical power to detect effects on expression. Comparison of the counts per million (CPM) obtained from DC-TAP-seq with the bulk TPM for the telo-HCASMC indicated good capture of most genes targeted in the PCR enrichment step (**Fig. 1B).** Each cell carried on average 3.28 gRNAs **(Fig. 1C, Suppl. Fig 2A-C)**, with an average of 555 transcripts per guide per cell. Using a simulation-based analysis of statistical power (Ray et al., in prep), 79% of tested element-gene pairs had >80% power to detect 25% effects on expression. (**Suppl. Fig. 2A, B**).

We identified 75 significant variant-to-enhancer-to-gene (V2E2G) links for 59 unique genes in 75 unique CAD loci (SCEPTRE ^22^, FDR < 0.1 (**Fig. 1D, E, Suppl. Fig. 2C-E, Table1, Suppl. Table 5)**. These V2E2G links have been uploaded to the Washington University Epigenome browser for visualization (https://shorturl.at/8P2qf). Most V2E2G links spanned <100 Kb from the perturbed element to the target gene promoter, and effect sizes on gene expression were inversely correlated with distance, similar to previous studies ^15,21^ (Ray et al. in prep) **(Fig. 1F).** Further characterization of the V2E2G links indicated that most significant genes (56) were associated with exactly one enhancer, and 9 genes were linked to 2 or 3 enhancers **(Fig. 1G).**

Multiple approaches were employed to validate these findings. The V2E2G links were compared to HiC interactions which demonstrated greater co-localization of HiC interactions than non-significant V2E2G links, irrespective of enhancer to gene distance (p<2.2e-16) **(Fig. 1H).** Further, we validated the DC-TAP-seq enhancer screen results by conducting CRISPRi-PCR in the HCASMC-CRISPRi cells, and 12 of the DC-TAP-seq identified links were validated in this fashion, including enhancers for *CKB, PPP1R18, RAB23, EDEM2, MICA, TP53INP2*, (**Fig. 1I-K, and Suppl. Fig. 2F-H)**. Six of the DC-TAP-seq genes have been validated as GWAS causal genes for CAD, including *MYO9B* ^2^, *FURIN* ^23^, *ADAMTS7* ^24^, *CDKN2B* ^25,26^*, COL4A1* ^27^, and *SMAD3* ^9^ **(Suppl. Fig 2I-K).** Thus, over 25% of the multi-locus screen identified genes have been substantiated as causal for CAD or validated as targets for enhancers residing in GWAS loci.

To further substantiate these V2E2G genes as relevant to CAD, we compared them to genes identified by polygenic priority score (PoPS) algorithm, which prioritizes causal genes through consideration of gene features, such as cell-type-specific expression or gene pathway information, independent of information about specific variants or enhancers in a given locus ^28^. We first intersected our 470 candidate genes with those predicted by PoPS and found that 88.7% were also identified with that approach **(Fig. 1L)**. For TAP-seq identified genes, 95% had positive PoPS scores. Comparison of the negative log p-values from Perturb-seq with scores derived from PoPS indicated significant overlap with the top PoPS gene, with UQCC2 being the top hit for both approaches. However, overall TAP-seq genes were a small percentage of the total genes identified with PoPS, and the overall correlation was low for this reason.

To benchmark the TAP-seq approach against additional genome wide studies in the field, we conducted a precision recall analysis to assess the ability of our Perturb-seq approach to identify eight gold standard SMC genes **(Fig. 1M)**. These gold standard genes included those identified through CAD GWAS, or identified as serving important functional roles in the SMC lineage, (*CDKN2B, TCF21, SMAD3, BMP1, LOXL1, MYO9B, PALLD, FN1*) ^2^. V2G2P performed well compared with studies that nominated CAD genes, including “locus to gene” (L2G) that employs a machine-learning model to learn the weights of evidence based on gold standard genes, as well as fine mapping and colocalization data ^29^. Also included in this analysis was the PoPS result, a transcriptome wide association study of CAD ^30^, a CAD GWAS meta-analysis and fine mapping study ^17^, and a CAD meta-analysis that reported eQTLs and sQTLs ^31^. Specifically, the TAP-seq approach achieved higher recall for the gold standard genes (50% compared to lower results for all others except PoPS) while also achieving high precision (43%). Beyond these 8 gold-standard genes, there was also significant enrichment of our identified CAD genes among previously prioritized lists of candidate CAD genes, generated by groups employing fine-mapping, Mendelian randomization, and colocalization approaches (e.g., FDR 1.68e-06 – 3.9e-10) ^32–35^.

To investigate commonality of transcriptional regulation at the V2E2G enhancers, we scanned transcription factor motifs in these significant enhancer regions, using non-significant enhancers as background **(Fig. 1N).** We identified primarily modest enrichment of AP-1 transcription factor binding motifs, as well as binding sites for ELK and ETS family motifs, and at a much lower level of enrichment motifs for SOX and GATA factors. These results suggest that multiple signaling pathways contribute to the regulation of these V2E2G causal gene networks.

### CAD causal genes identify a variety of CAD cellular programs in transition SMC

Inspection of the biological and molecular functions of the 59 identified CAD genes identified a broad spectrum of biological pathways represented among the identified gene functions **(Table 1).** By allowing the STRING algorithm to incorporate a small number of additional Perturb-Seq identified genes that mediated protein-protein interactions (PPIs) with the members of the CAD TAP-seq genes, it was possible to elucidate a CAD gene network that was enriched for numerous biological processes. This network showed enrichment for terms related to “protein folding,” “ubiquitination,” “cytoskeleton,” “GTPase function,” and “BMP” and “TGFB functions” **(Fig. 2A, B).** The added seed genes did not represent a majority of contributors to any of the identified pathways and by themselves did not identify significant gene ontology terms, suggesting that they strengthened the signal for gene sets that would not otherwise be identified in the context of the large number of disparate biological processes represented by the V2E2G genes. Gene set enrichment analysis of this combined PPI geneset also identified numerous terms related to osteochondrognic processes, including “regulation of cartilage development” and “ossification,” reflecting the SMC transition to the chondromyocyte phenotype, as well as extracellular matrix terms **(Fig. 2B)**.

**Table 1.**
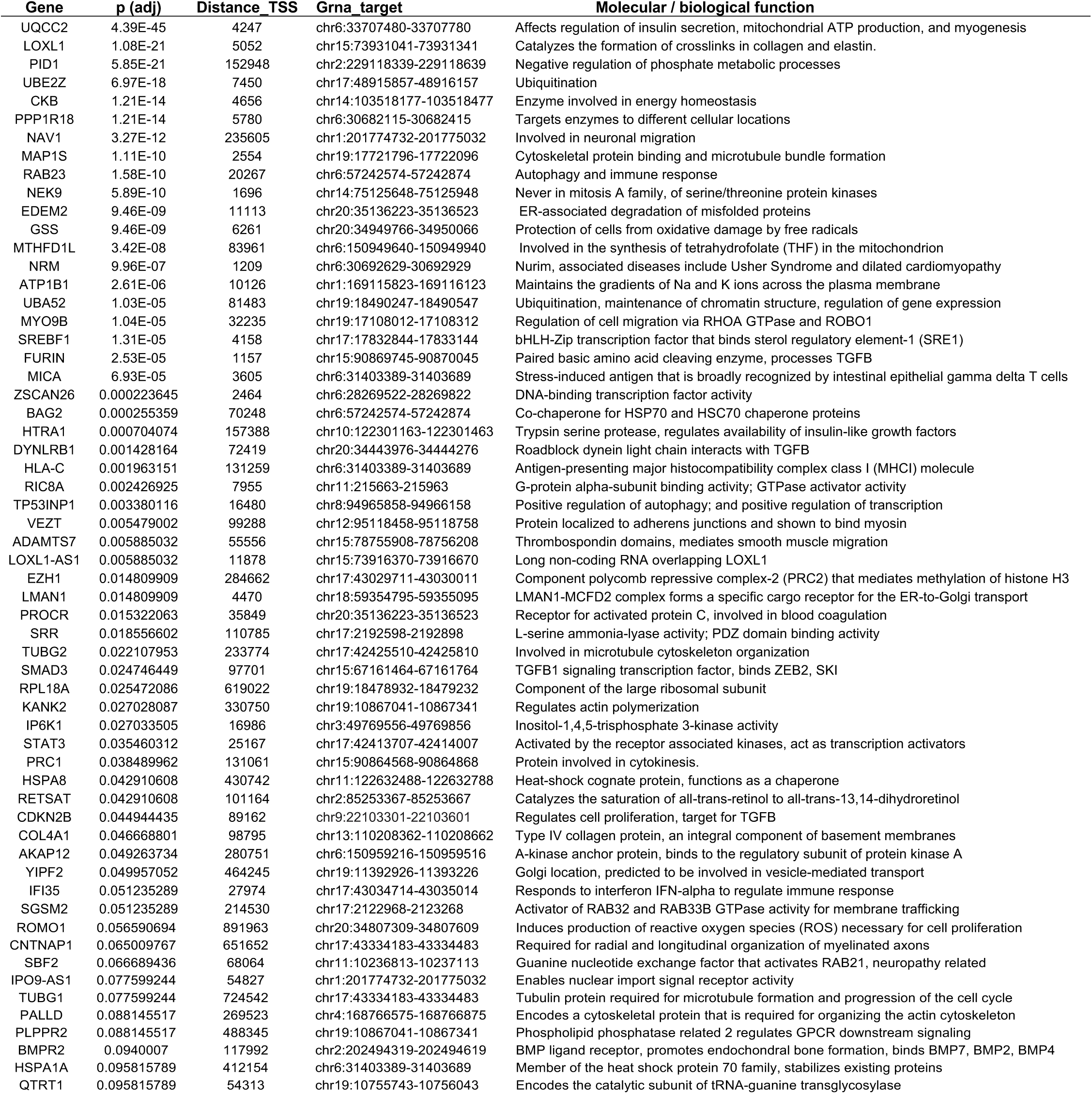
Candidate CAD causal genes identified by multi-locus CRISPRi screen.

**Figure 2.**
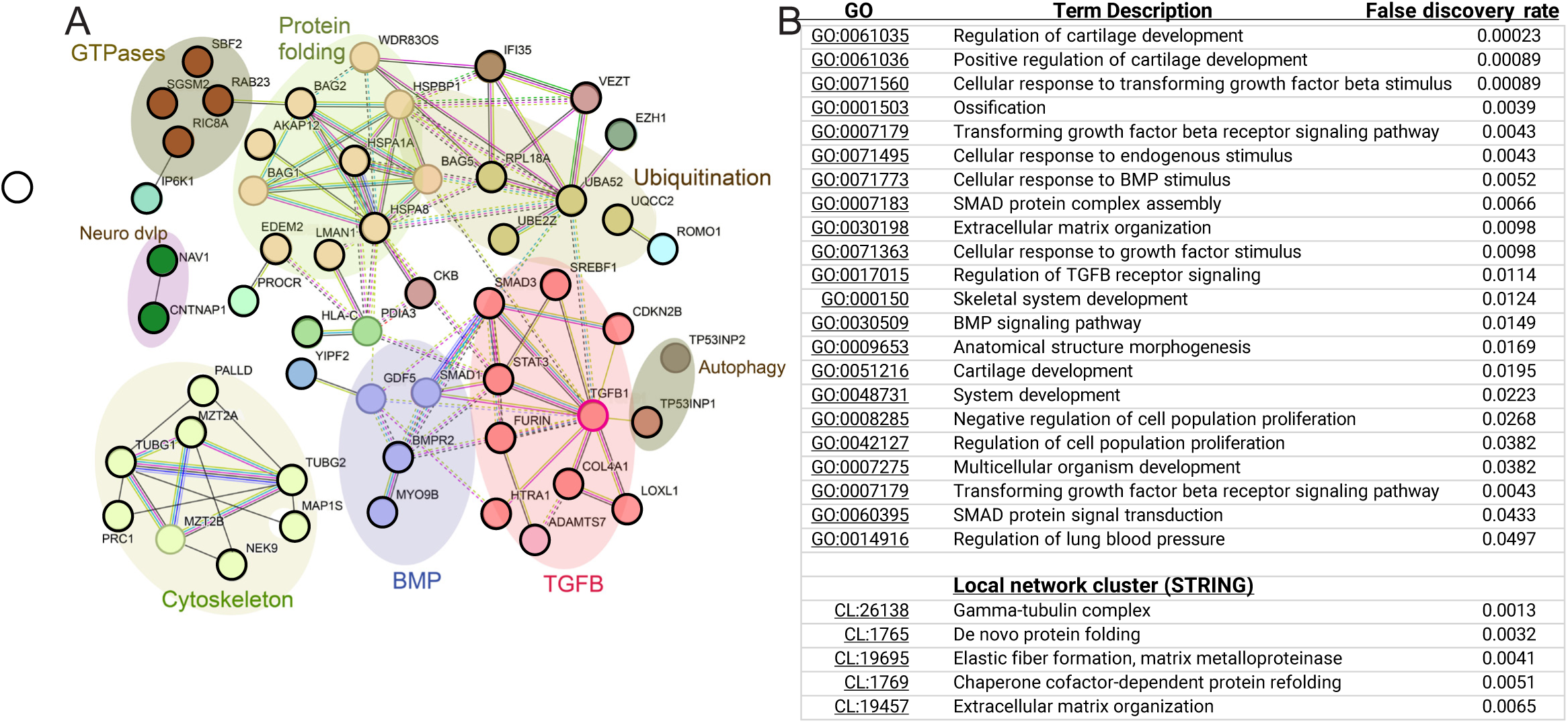
CAD GWAS enhancer target genes identified with CRISPRi screen map to functional gene programs. **A)** Total 59 unique target genes showed numerous interconnections and related functions. Nodes with black rings indicate genes identified by the multi-locus DC-TAP-seq, while genes added by STRING do not have this marking. Genes with similar color are clustered by Markov Chain method and are enriched in functional pathways. **B)** Functional gene programs identified with the STRING protein-protein interaction database. Gene Ontology terms, gene annotation, and Gene Set Enrichment Analysis false discovery rate are shown.

### DC-TAP-seq at 9p21.3 reveals numerous individual SMC enhancers that mediate V2E2G links

One enhancer identified in the DC-TAP-seq experiment, located at the 9p21.3 locus, was found to regulate the expression of CAD candidate gene *CDKN2B*. Given that this GWAS locus contributes a significant portion of CAD risk in the human genome, with variation observed across sexes and ancestry groups, we pursued additional studies to examine mechanisms of CAD risk at this locus ^3,36–38^. Therefore, we designed an additional DC-TAP-seq screen to perturb candidate enhancers in this locus **(Fig. 3A).** We used LD expanded 9p21.3 CAD GWAS variants and intersected them with epigenetic data from three datasets: scATACseq data from telo-HCASMC ^5^, bulk ATACseq and bulk H3K27ac obtained from the telo-HCASMC. This identified 10 enhancers containing 27 SNPs **(Fig. 3A, B**, **Table 2, Suppl. Table 6).** Guide RNAs were designed as described for the multi-locus screen with the CRISPRi-dCas9-mediated direct capture vector approach, employing a dCas9-KRAB-ZIM3 transcriptional inhibitor. These enhancer elements were targeted in CRISPRi-HCASMC with 200 gRNAs **(Suppl. Table 6)** and identified candidate genes with DC-TAP-seq amplification of 23 genes, including 7 in the 9p21.3 locus and the rest positive and negative controls **(Suppl. Table 7**). Results for 4 genes in the 9p21.3 locus are shown **(Table 2)**. Power analyses indicated that we had very high power (>95%) to detect 25% effects on expression for 5 of 7 genes in the locus (*CDKN2A, CDKN2B, IFNE, KLHL9*, and *MTAP*), and had much lower power for the remaining 2 genes (*DMRTA1* and *CDKN2B-AS1*), which were very lowly expressed (**Suppl. Fig. 3A, B)**.

**Figure 3.**
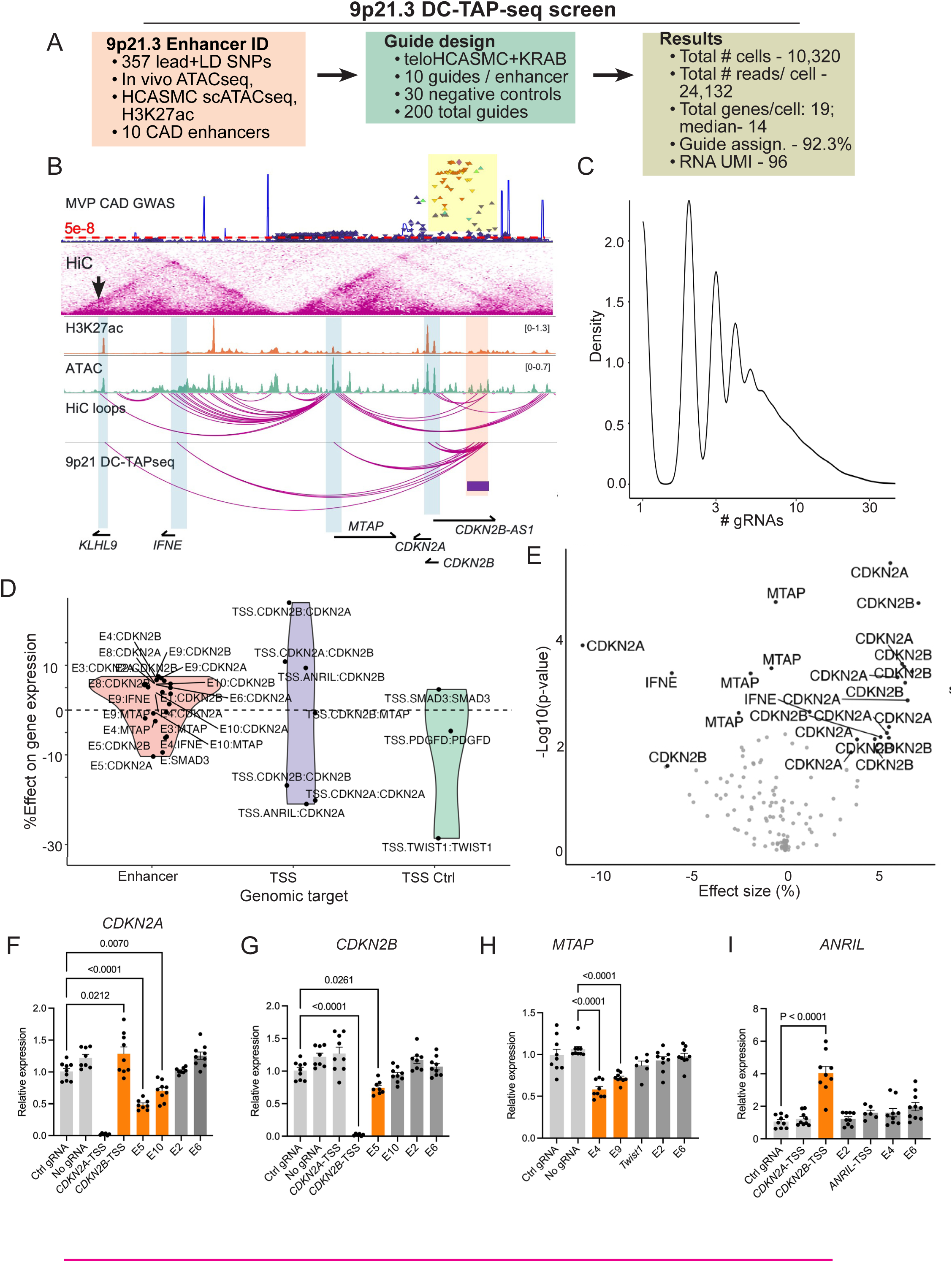
CRISPRi screen for mapping of CAD GWAS enhancers to candidate causal genes at 9p21.3. **A)** Design of SMC in vitro DC-TAP-seq targeting screen of CAD enhancers at 9p21.3. **B)** Details regarding CAD GWAS enhancer identification, gRNA design for CRISPRi epigenome editing, and screen results. **C)** Graphical representation of guide RNAs (gRNAs, protospacers) transduced into CRISPRi-HCASMC by lentiviral infection. **D)** Violin plot showing effect of gRNA directed gene knockdown and resulting effects on gene expression. Experimental conditions were gRNA directed to CAD GWAS enhancers, targeting of TSS for 9p21.3 experimental genes, and targeting of TSS for known CAD causal genes *PDGFD* and *SMAD3*. **E)** Volcano plot showing effect on gene expression of gRNA targeted enhancers identified at 9p21.3, graphed as log fold change versus effect size. Grey dots represent genes not affected by enhancer knockdown, red dots and gene symbols represent genes showing significant effect toward gRNA targeting, average of multiple guides targeted to specific sequences in the 9p21.3 enhancers. **F)** Genome browser image showing long range chromatin organization at this locus, H3K27ac histone modification from ChIPseq, bulk ATACseq, and HiC loops all mapped in telo-HCASMC. Also, shown are genes encoded in this locus, and the E2G links identified by DC-TAP-seq. **G, H)** CRISPRi epigenome editing in CRISPRi-HCASMC transduced with lentivirus control gRNAs, no gRNAs, *CDKN2A* transcription start site (TSS), *CDKN2B* TSS, and sgRNAs for enhancers expected to be positive or negative. **I)** A similar PCR quantitation of *MTAP* expression was performed with CRISPRi-HCASMC transduced with lentivirus control gRNAs, no gRNAs, and gRNAs targeting the TSS of CAD *Twist1* as a positive control, and sgRNAs for enhancers expected to be positive or negative. **J)** Enhancer control of *ANRIL* expression was investigated with gRNAs targeting E2, E4, and E6. Experiments were conducted with *CDKN2A* and *CDKN2B* TSS gRNAs to investigate gene crosstalk.

**Table 2.**
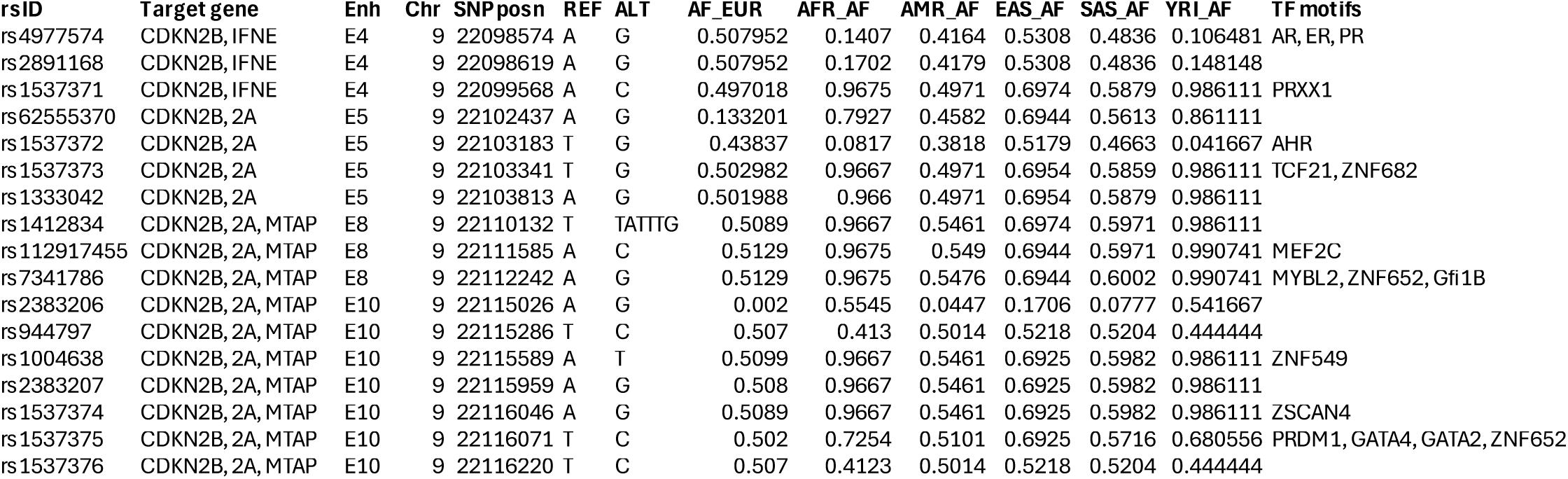
9p21.3 enhancers, target genes, and related CAD associated variants and their allele frequencies across different ancestry groups. These variants represent those investigated for epigenetic and transcriptional functional activity toward genes in this locus.

| rsID | Target gene | Enh | Chr | SNPposn | REF | ALT | AF_EUR | AFR_AF | AMR_AF | EAS_AF | SAS_AF | YRI_AF | TF motifs |
| --- | --- | --- | --- | --- | --- | --- | --- | --- | --- | --- | --- | --- | --- |
| rs4977574 | CDKN2B, IFNE | E4 | 9 | 22098574 | A | G | 0.507952 | 0.1407 | 0.4164 | 0.5308 | 0.4836 | 0.106481 | AR, ER, PR |
| rs2891168 | CDKN2B, IFNE | E4 | 9 | 22098619 | A | G | 0.507952 | 0.1702 | 0.4179 | 0.5308 | 0.4836 | 0.148148 |  |
| rs1537371 | CDKN2B, IFNE | E4 | 9 | 22099568 | A | C | 0.497018 | 0.9675 | 0.4971 | 0.6974 | 0.5879 | 0.986111 | PRXX1 |
| rs62555370 | CDKN2B, 2A | E5 | 9 | 22102437 | A | G | 0.133201 | 0.7927 | 0.4582 | 0.6944 | 0.5613 | 0.861111 |  |
| rs1537372 | CDKN2B, 2A | E5 | 9 | 22103183 | T | G | 0.43837 | 0.0817 | 0.3818 | 0.5179 | 0.4663 | 0.041667 | AHR |
| rs1537373 | CDKN2B, 2A | E5 | 9 | 22103341 | T | G | 0.502982 | 0.9667 | 0.4971 | 0.6954 | 0.5859 | 0.986111 | TCF21, ZNF682 |
| rs1333042 | CDKN2B, 2A | E5 | 9 | 22103813 | A | G | 0.501988 | 0.966 | 0.4971 | 0.6954 | 0.5879 | 0.986111 |  |
| rs1412834 | CDKN2B, 2A, MTAP | E8 | 9 | 22110132 | T | TATTG | 0.5089 | 0.9667 | 0.5461 | 0.6974 | 0.5971 | 0.986111 |  |
| rs112917455 | CDKN2B, 2A, MTAP | E8 | 9 | 22111585 | A | C | 0.5129 | 0.9675 | 0.549 | 0.6944 | 0.5971 | 0.990741 | MEF2C |
| rs7341786 | CDKN2B, 2A, MTAP | E8 | 9 | 22112242 | A | G | 0.5129 | 0.9675 | 0.5476 | 0.6944 | 0.6002 | 0.990741 | MYBL2, ZNF652, Gfi1B |
| rs2383206 | CDKN2B, 2A, MTAP | E10 | 9 | 22115026 | A | G | 0.002 | 0.5545 | 0.0447 | 0.1706 | 0.0777 | 0.541667 |  |
| rs944797 | CDKN2B, 2A, MTAP | E10 | 9 | 22115286 | T | C | 0.507 | 0.413 | 0.5014 | 0.5218 | 0.5204 | 0.444444 |  |
| rs1004638 | CDKN2B, 2A, MTAP | E10 | 9 | 22115589 | A | T | 0.5099 | 0.9667 | 0.5461 | 0.6925 | 0.5982 | 0.986111 | ZNF549 |
| rs2383207 | CDKN2B, 2A, MTAP | E10 | 9 | 22115959 | A | G | 0.508 | 0.9667 | 0.5461 | 0.6925 | 0.5982 | 0.986111 |  |
| rs1537374 | CDKN2B, 2A, MTAP | E10 | 9 | 22116046 | A | G | 0.5089 | 0.9667 | 0.5461 | 0.6925 | 0.5982 | 0.986111 | ZSCAN4 |
| rs1537375 | CDKN2B, 2A, MTAP | E10 | 9 | 22116071 | T | C | 0.502 | 0.7254 | 0.5101 | 0.6925 | 0.5716 | 0.680556 | PRDM1, GATA4, GATA2, ZNF652 |
| rs1537376 | CDKN2B, 2A, MTAP | E10 | 9 | 22116220 | T | C | 0.507 | 0.4123 | 0.5014 | 0.5218 | 0.5204 | 0.444444 |  |

Analysis of gRNA sequencing in captured cells indicated that less than 10% of cells had no guide identified, less than one-fourth had one assigned gRNA, and more than three-fourths had more than one guide with a declining distribution from one to five gRNAs per cell **(Fig. 3C).** As for the multi-locus screen, we analyzed the resulting data with SCEPTRE, which had difficulty with calibration of this small dataset, but did identify *CDKN2B* (adjusted p=3.67e-5) and *CDKN2A* (adjusted p=2.67e-4) as target genes for enhancers 4 and 5 respectively **(Suppl. Table 8).** As an alternative analysis approach, we employed the two-part generalized linear model implemented in the MAST algorithm by Finak et al, to assess differential gene expression in single cell data **(Suppl. Table 8)** ^16,39^. This analytical approach identified four gene targets and was bolstered with extensive validation and mechanistic experimentation.

MAST identified multiple significant enhancer-to-gene links at the 9p21 locus, and these links matched HiC loops identified in HCASMC (**Fig. 3B, D, E, Suppl. Fig. 3C, Suppl. Fig. 4A, C, Table 2, Suppl. Table 9**). V2E2G connections were identified for several of the 10 enhancers showing regulation of *CDKN2A, CDKN2B,* and *MTAP*. In addition to these three genes, a number of the guides showed knockdown for *IFNE,* which encodes a type I interferon that regulates expression of ISG15 and other anti-viral and -bacterial proteins. Interestingly, CRISPRi epigenetic editing of these genes by different enhancers had differing directions of effect. For instance gRNA directed targeting of E5 led to a decrease in expression of *CDKN2A* while targeting of E8 led to increased expression of this gene **(Fig. 3D)**. Also guides targeting the same enhancer were found to have opposite effects on different genes, with E4 inhibition promoting decreased expression of *IFNE* and increased expression of *CDKN2B*. These differential effects may be explained by the use of different gRNAs targeting distinct nearby enhancers, or by variability in enhancer function, where some enhancers may exert activating or repressive effects depending on the context of the target gene’s promoter ^40^. Some guides showed an effect toward knocking down expression of *KHLH9*, although the results were not consistent enough among the guides to show an overall significant effect. HiC data generated previously in this lab ^19^ identified looping between *KHLH9,* and the enhancer region identified through these studies **(Fig. 3B)**. The function of *KHLH9* as a critical mediator of cell division would be consistent with the function of the CDKIs in this locus.

In this screen we did not observe significant E2G effects for the long non-coding gene *ANRIL* (*CDKN2B-AS1*) which has been linked to multiple disease related features and CAD risk through function at this locus ^41–44^. This could be due to the low power for study of this gene and its low expression level. However, targeting the TSS of *ANRIL* produced a decrease in *CDKN2A* and an increase in *CDKN2B*, suggesting that it plays a functional role in regulating the expression of both these genes **(Fig. 3D)**. These effects are unlikely to be off-target effects of the *ANRIL* promoter gRNAs toward promoters of these genes as they are over 10 kb from the *ANRIL* promoter. The *ANRIL* effect on *CDKN2B* expression could be due to the known transcriptional inhibition of *CDKN2B* through an antisense effect as these genes are overlapping and transcribed in the opposite direction ^45^. It is also worth noting that targeting the TSS of each of the CDKI genes led to increased expression of the other **(Fig. 3D)**. Such counter-regulatory effects of CDKI expression in this locus have been previously described ^26,45–47^.

Finally, we validated the 9p21.3 DC-TAP-seq results using individual single gRNAs with CRISPRi and CRISPRa editing with the same gRNAs and individual qPCR reactions. Although not all E2G links identified with the locus-wide screen were confirmed, these assays validated E5 and E10 regulation of *CDKN2A*, E5 regulation of *CDKN2B*, and E4 and E9 regulation of *MTAP* **(Fig. 3F-H)**. CRISPRi knockdown of E2, E4 and E6 did not show a discernible decrease in lowly expressed lncRNA *ANRIL* expression, and indeed gRNAs targeting the *ANRIL* TSS also failed to produce decreased expression **(Fig. 3I).** Similar CRISPRa studies in HCASMC expressing a dCas9-VPR editor did show that activation of all 10 enhancers combined significantly upregulated *ANRIL* expression, and activation of E5 also nominally upregulated *ANRIL* expression (p=0.058) **(Suppl. Fig. 4C)**. The CRISPRa studies further showed that activation of all 10 enhancers upregulated *CDKN2A, CDKN2B*, and *MTAP*, with E5 activation also increasing *CDKN2A* expression **(Suppl. Fig. 4D-F)**. As positive controls, we had included gRNAs designed to target the TSS for the 9p21 genes, and these experiments provided further evidence for counter-regulation between the different genes at this region of the human genome. TSS targeting of *CDKN2B* resulted in upregulation of *CDKN2A* **(Fig. 3F)** and targeting of *CDKN2B* produced an increase in *ANRIL* expression **(Fig. 3I).**

### Causal variants at 9p21.3 enhancers mediate ancestry- and sex-specific CAD risk

9p21.3 is one of only a small number of loci where allelic variation has been associated with differences in CAD risk attributed to ancestral genetic background ^3^ or sex ^36^, or identified as highly associated with clinical vascular calcification ^37^. Given that we had mapped the variants and enhancers that were likely responsible for disease association at this locus, we were interested to determine the mechanisms by which they might contribute to these characteristics of local CAD risk. Therefore, we mapped all the CAD associated lead variants and their associated proxies at LD r^2^≥0.8, that reside within 1 kilobase of the gRNA targeted areas. We identified five SNPs in E5, three SNPs each in E4 and E8, and seven SNPs in E10. We gathered allele frequencies for all these variants across ancestries and highlighted the ones which vary across ancestry, sex, and were associated with vascular calcification (**Table 2)**. Next, we used ChromBPNet variant scoring ^48^, Jaspar ^49^, Hocomoco ^50^ and TomTom ^51^ resources to identify transcription factor (TF) binding motifs that overlapped CAD associated variants in our identified enhancers, and we prioritized further studies for the variants predicted to regulate chromatin accessibility or disrupt TF binding.

#### Aryl hydrocarbon receptor (AHR) binding, transcriptional regulation, and CAD risk mediated at rs1537372

Colocalization of CAD variants and TF motifs identified variant rs1537372 as possibly regulating TF binding at enhancer E5. This variant was highly significantly associated with CAD in the recent Million Veterans Study, association p=1.7e-309 compared to p=4.94e-324 for lead SNP rs10738610 in that study ^3^. This variant was also the lead SNP in a GWAS study for CAD risk in Taiwanese (^52^) and a GWAS study for carotid calcification ^53^. It was further noted to be in high LD with lead variant rs10738610 found to be significantly associated with severity of CAD in subjects with type 2 diabetes, r2 = 0.8 in TopLD ^54^.This variant resides in a motif for the AHR TF, and colocalizes with ChIPseq peaks for AHR and its obligate heterodimer ARNT, in prominent H3K4me1 and H3K27ac histone modification and scATACseq peaks, all mapped in HCASMC **(Fig. 4A).** The AHR binding motif is highly conserved across evolution, and the rs1537372 variant position lies at a critical location in this motif ^55,56^. Therefore, we suspected this variant would disrupt binding of AHR at this genomic site and impair transcriptional function of this region of E5.

**Figure 4.**
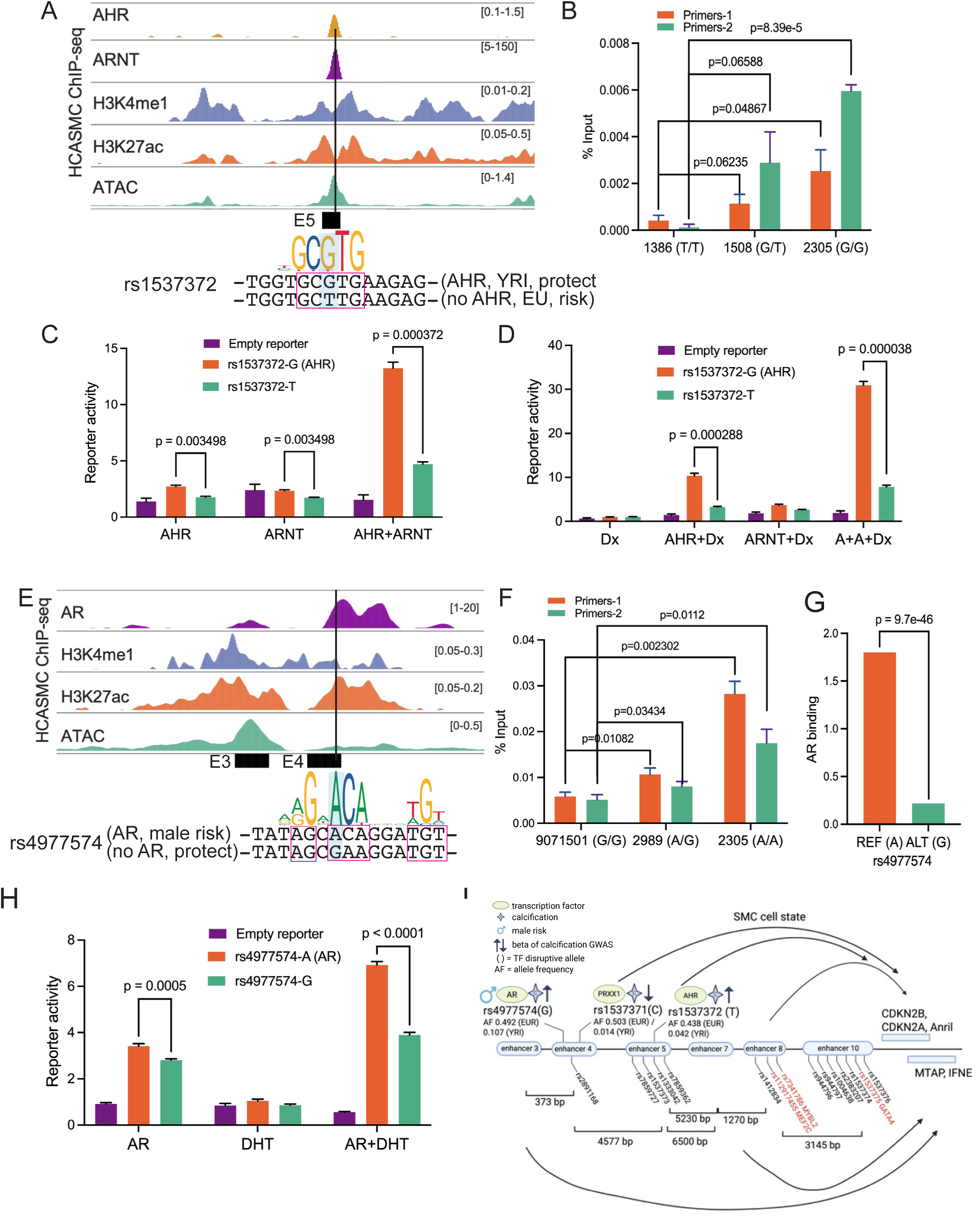
Candidate causal variants regulate ancestry- and sex-specific enhancer functions at 9p21.3. **A)** CAD variant rs1537372 and aryl hydrocarbon receptor (AHR) transcription factor binding motif co-localize with AHR, AHR binding partner ARNT, H3K4me1, and H3K27ac ChIP-seq peaks and a bulk ATACseq peak, all identified in HCASMC. rs1537372 alt T allele is predicted to disrupt the motif and transcription factor binding. **B)** ChIP-qPCR using two different AHR binding site primer pairs and HCASMC lines with differing genotypes at rs1537372 shows greater DNA immunoprecipitation and amplification in a G-allele specific pattern. **C)** AHR and ARNT transfection studies with reporter genes encoding rs1537372-G and -T alleles were evaluated in HEK293 cells. AHR and ARNT transfection shows a modest but significant increase in reporter gene transcription with the reference G-allele, and the combination of both transcription factors show a greater and more significant increase in reporter activity. **D)** Transfection of AHR and AHR plus ARNT in the context of AHR activation by 10nM dioxin showed greater reporter activity with the reference G-allele reporter construct in HEK293 cells. **E)** CAD variant rs4977574 and the androgen receptor (AR) transcription factor binding motif co-localize with AR, H3K4me1, and H3K27ac ChIP-seq peaks and a minimal bulk ATACseq peak, all identified in HCASMC. rs4977574 alt G allele is predicted to disrupt the motif and transcription factor binding. **F)** ChIP-qPCR using two different AR binding site primer pairs and HCASMC lines with differing genotypes at rs4977574 shows greater DNA immunoprecipitation and amplification in an A-allele specific pattern. **G)** Analysis of published motif-resolution chromatin immunoprecipitation-exonuclease (ChIP-exo) data (GSE43785) from studies in prostate cell lines revealed a significant allelic imbalance with greater AR binding to the A-reference allele compared to the G-alt allele ^65^. **H)** AR transfection studies with reporter genes encoding rs4977574-A and -G alleles were evaluated in HEK293 cells. AR transfection alone showed an increase in reporter gene transcription for both alleles, but transcription of the A allele was significantly greater. Addition of the 0.23nM androgen hormone dihydrotestosterone produced an increase for both allele reporters, but activity of the A-reporter was even greater than activation of the G-reporter allele. **I)** Annotation of the enhancers and variants that function at the 9p21.3 locus to contribute to expression of genes in this locus and the risk for CAD.

We also noted that this variant showed significant variability with respect to allele frequencies across different ancestral genetic backgrounds. Individuals of northern European, East Asian, South Asian, and admixed African American ancestry had reference A allele frequencies of ∼0.4 to ∼0.5, while individuals of mixed African ancestry had A allele frequencies of 0.08 and most notably those of Yoruba descent had approximately 10-fold lower A allele frequencies of ∼0.04 **(Table 2, Suppl. Table 9)**. The YRI allele was not in linkage disequilibrium with any other alleles in the YRI haplotype as represented in 1000 Genomes and TopLD **(Suppl. Fig. 5A).** It has been shown previously that individuals of African ancestry have a lower level of CAD risk at 9p21 ^3^. Also, the haplotype structure at 9p21.3 has been described in relation to CAD risk, providing a roadmap for studies investigating the relationship between rs1537372 genotype, AHR binding, transcriptional regulation, and gene expression at this locus ^3^. We thus performed a genotype analysis at this locus to identify HCASMC lines carrying the African Yoruba (YRI) and European (EUR) haplotypes on one or both alleles. To determine local ancestry, we used FLARE (Fast Local Ancestry Estimation ^57^ to map variants from whole-genome sequencing data on chromosome 9 to the 1000 Genomes reference dataset with population annotations ^58^. The population to which more than 90% of the variants were mapped was considered the ancestral origin. To investigate AHR binding at this variant, relevant lines were used to perform ChIP-PCR studies with an AHR antibody and chromatin isolated from HCASMC that were homozygous for the reference allele (G/G), heterozygous for the reference and alternative alleles (G/T) or homozygous for the alternative alleles (T/T) **(Fig. 4B, Suppl. Table 10)**. These experiments, employing two different PCR primer sets, and #1386 EUR T/T, #1508 EUR G/T, and #2305 AMR G/G genotype lines, showed a dose-dependent highly significant G-allele specific binding of AHR.

Further studies were conducted to investigate transcriptional activity mediated by AHR binding to the two alleles, and whether AHR bound to these alleles can respond to transcriptional activation by the AHR ligand dioxin. For these studies we employed genomic sequence blocks amplified from the homozygous TT and GG HCASMC and thus representing the independent T and G alleles, in the context of reporter gene studies. These genomic blocks were cloned into a minimal luciferase reporter transfected into human HEK 293 kidney cells and rat aortic A7R5 SMC and transcriptional activity determined with dual luciferase-renilla assays **(Fig. 4C, Suppl. Fig. 5B, Suppl. Table 11).** Expression vectors encoding AHR and its heterodimer partner ARNT were co-transfected and assayed separately and combined. Increased expression of both AHR and ARNT individually and combined provided significantly greater reporter expression of the AHR binding rs1537372 G-allele, compared to the minor T-allele, with greater overall and differential expression when both AHR and ARNT vectors were transfected. AHR showed greater transcriptional activity when it binds the chemical dioxin (2,3,7,8-tetrachlorodibenzo para-dioxin, TCDD) ^59,60^, and thus the dioxin stimulated AHR effect on transcription mediated by the two alleles was evaluated **(Fig. 4D, Suppl. Fig. 5B, Suppl. Table 11).** These studies showed increased transcriptional activity for the both the AHR binding G-allele and the T-allele in the dioxin stimulated condition, but with a 3.5-fold greater increase for the G-allele containing reporter vector.

While AHR has been studied as a mediator of environmental toxic pollutants such as dioxin, it plays a critical role in embryonic vascular development and other beneficial roles ^59,60^. Further, studies conducted in this lab have shown this gene to retard atherosclerosis development and vascular calcification in mouse disease models ^56^. In the context of these mouse model studies we analyzed previous scRNAseq data generated with diseased proximal aortic tissue, and found that knockout of A*hr* resulted in a highly significant increase in the expression of 9p21.3 genes *Cdkn2b* and *Cdkn2a*, p<300 for each. **(Suppl. Fig. 5C, D)**. This work did not indicate whether these effects were directly related to Ahr binding and transcriptional regulation, or through indirect effects, but they do indicate that regulation of these 9p21.3 genes is a component of the Ahr gene program, and consistent with the protective effect that the reference G-allele shows toward CAD risk in individuals with YRI haplotype alleles.

#### Androgen receptor (AR) binding, transcriptional regulation, and CAD risk mediated at rs4977574

Colocalization of CAD variants and TF motifs identified variant rs4977574 as possibly regulating TF binding at enhancer E4. This variant was identified as a lead SNP in multiple ancestry population GWAS and meta-analyses for CAD ^61–64^, and noted to be in high LD with the MVP lead variant rs10738610 in TopLD, (r2=0.95) ^3^. Variant rs4977574 was identified in a motif in enhancer E4 expected to bind hormone receptors testosterone, progesterone, or estrogen (**Fig. 4E, Suppl. Fig. 6A)**.

As for the AHR motif, this SNP constitutes an A/G basepair in a highly conserved location in the position weight matrix for hormone receptor TFs. This variant in the E4 enhancer was found to be located in a peak for AR binding as identified with an exonuclease (ChIP-exo) assay in human prostate cancer cells (GSE43785) ^65^. There was also colocalization for H3K4me1 and H3K27ac enhancer histone marks identified by ChIPseq in HCASMC, and modest chromatin accessibility as determined by ATACseq assayed in HCASMC. The alternative causal risk G allele has been associated with increased risk of CAD in men compared to women ^36,61^ and the DNA sequence at rs4977574 showed the greatest overlap with the AR binding motif. We were thus interested to determine whether the androgen hormone receptor might bind and functionally interact with this canonical binding site in a haplotype-specific fashion, and initial studies focused on this member of the hormone receptor family **(Fig. 4E-I, Suppl. Fig. 6B).**

We first investigated allele-specific binding of AR at the rs4977574 variant with ChIP-PCR studies employing an antibody to human AR and HCASMC lines with informative haplotypes for this variant and motif **(Fig. 4F, Suppl. Table 10)**. HCASMC studied were homozygous for the alternative allele (G/G), heterozygous for the reference and alternative alleles (A/G) or homozygous for the alternative allele (A/A). These experiments, employing two different PCR primer sets, and 9071501 EUR G/G, 2989 EUR G/T, and 2305 AMR A/A lines showed a dose-dependent highly significant G-allele specific binding of AR. We also identified allele-specific chromatin immunoprecipitation-exonuclease (ChIP-exo) studies that had been performed for AR in prostate cancer cells, and reported previously ^65^. Analysis of these data identified a relative lower level of binding to the G compared to the AR binding A allele **(Fig. 4G)**. Further, reporter gene transfection studies in HEK **(Fig. 4H)** and A7R5 SMC **(Suppl. Fig. 6B)** with an AR expression vector and reporter constructs containing the AR motif ref A-allele or the alt G-allele showed highly significant greater transcription of the A-allele, and stimulation of the cells with dihydrotestosterone (DHT) in the presence of AR provided significant overall greater levels of transcription **(Fig. 4H, Suppl. Fig. 6B, Suppl. Table 11).** The risk G-allele thus disrupts AR binding and provides for lower expression of the 9p21.3 genes, offering a mechanistic link for sex-biased CAD risk at this locus. This finding that AR function in SMC inhibits disease processes in this cell type, is consistent with evidence in the literature suggesting that AR signaling may be protective toward vascular cell disease related processes ^66,67^.

#### Genome-wide co-localization of AHR and AR binding and the functional interactions of these TFs in SMC transcription

Interestingly,rs1537372 and rs4977574 are in linkage disequilibrium (LD 0.80 in TopMed), suggesting that they function as a haplotype. Also, interaction between the AHR and AR signaling pathways has been known for some time, and these two TFs shown to physically interact ^68–71^.To assess whether AHR and AR bind in juxtaposition across the genome, and thus may interact through cooperative regulation of nearby genes, we investigated their co-localization across the genome. We utilized ChIP-seq data for the two transcription factors. For AR we employed the GSE268252 ChIP-exo data described above for AR binding in prostate cancer cells ^72^, and compared these data to ChIP-seq data generated in this lab for AHR binding in HCASMC (GSE276152) ^73^. Mapping of AHR binding sites to AR sites showed highly significant enrichment of AHR binding nearby to AR binding sites (p <2.2e-16) by the Wilcoxon rank sum test) **(Suppl. Fig. 7A).**

To further investigate a relationship at the genetic level, and generalize the findings at 9p21.3 to other CAD loci, we investigated the genome-wide occurrence of predicted motif disruptions in AHR and AR motifs in CAD loci. We first identified CAD GWAS SNPs by expanding the linkage disequilibrium (LD) (r^2^≥0.8) of the initial 357 lead SNPs ^3^, yielding a total of 8,395 SNPs. We then extracted genomic sequences surrounding these SNPs based on the analyzed motif lengths. For AR and its negative control, HNF1A, we included ±15 nucleotides around each SNP, while for AHR/ARNT and the negative controls, GATA1 and ZNF354C, we included ±6 nucleotides to align with their basic motif lengths. Each sequence was generated in two versions, incorporating either the reference or alternative allele according to dbSNP151. These sequences were then used to compute PWM probability scores of the motifs using TFBStools. The differences in scores between the reference and alternative alleles were analyzed to evaluate the impact of SNPs on transcription factor binding potential **(Suppl. Fig. 7B, C).** For AR, we identified enrichment of CAD associated variants located in TF motifs that were predicted to disrupt binding (p <2.2e-16 by the Wilcoxon rank sum test). The distribution of score differences showed greater perturbation of AR than the control TF HNF1A. For AHR, there was again evidence for enrichment of CAD variants among those that were predicted to have a detrimental effect on TF binding, equally significant to the AR analysis, but the distribution of differences was not different than seen for the control TFs.

Given the evidence for direct AHR-AR protein binding, and evidence that AHR can regulate AR signaling ^68,70^, we investigated the functional interaction of these TFs toward transcriptional regulation at the rs49777547 AR site in SMC. A luciferase reporter plasmid containing amplified DNA homozygous for the AR binding site at rs4977574 (A allele) was transfected into SMC with expression vectors encoding AHR, ARNT, AR, or all three TFs. AR alone promoted significantly increased transcription while AHR and partner ARNT had negative or modest increased transcriptional effects **(Fig. 7D)**. The combination of all three TFs showed even greater promotion of transcription which was further increased by the stimulation of transfected cells with DHT.

These studies point to relevance of direct AHR regulation of disease risk, the ability of AHR to locally mediate the transcriptional functions and thus risk due to AR binding, and the possibility for AHR being responsible for mediating CAD risk by mediating the effects of environmental factors.

### 9p21.3 CAD enhancers regulate SMC cell state

We have identified CAD associated enhancers that regulate expression of 9p21.3 encoded genes, and the V2E2G links that mediate some of these effects. To verify that these enhancers can modulate cell state changes that might be regulated through these enhancers and genes, we have performed in vitro studies with CRISPRi-HCASMC expressing dCAS9-KRAB-ZIM3 modeling enhancer suppression. Cellular proliferation studies employed HCASMC with two YRI, two EUR, and EUR alleles on a Hispanic ethnic background (EUR-Hispanic). Cellular proliferation was investigated by EdU staining and a fluorescent-activated cell sorting assay **(Fig. 5A)**. When a single gRNA expressing lentivirus targeting E5 was transduced into EUR cells, or a mixture of viruses encoding all ten enhancers was transduced, these assays showed a significant increase in cellular proliferation. The EUR-Hispanic cells were also able to show increased cell division in the mixed gRNA condition. Interestingly, using gRNAs targeting individual enhancers E4 or E5 or all the enhancers identified at this locus, we did not detect a difference in rate of cell division for the YRI allele cells.

**Figure 5.**
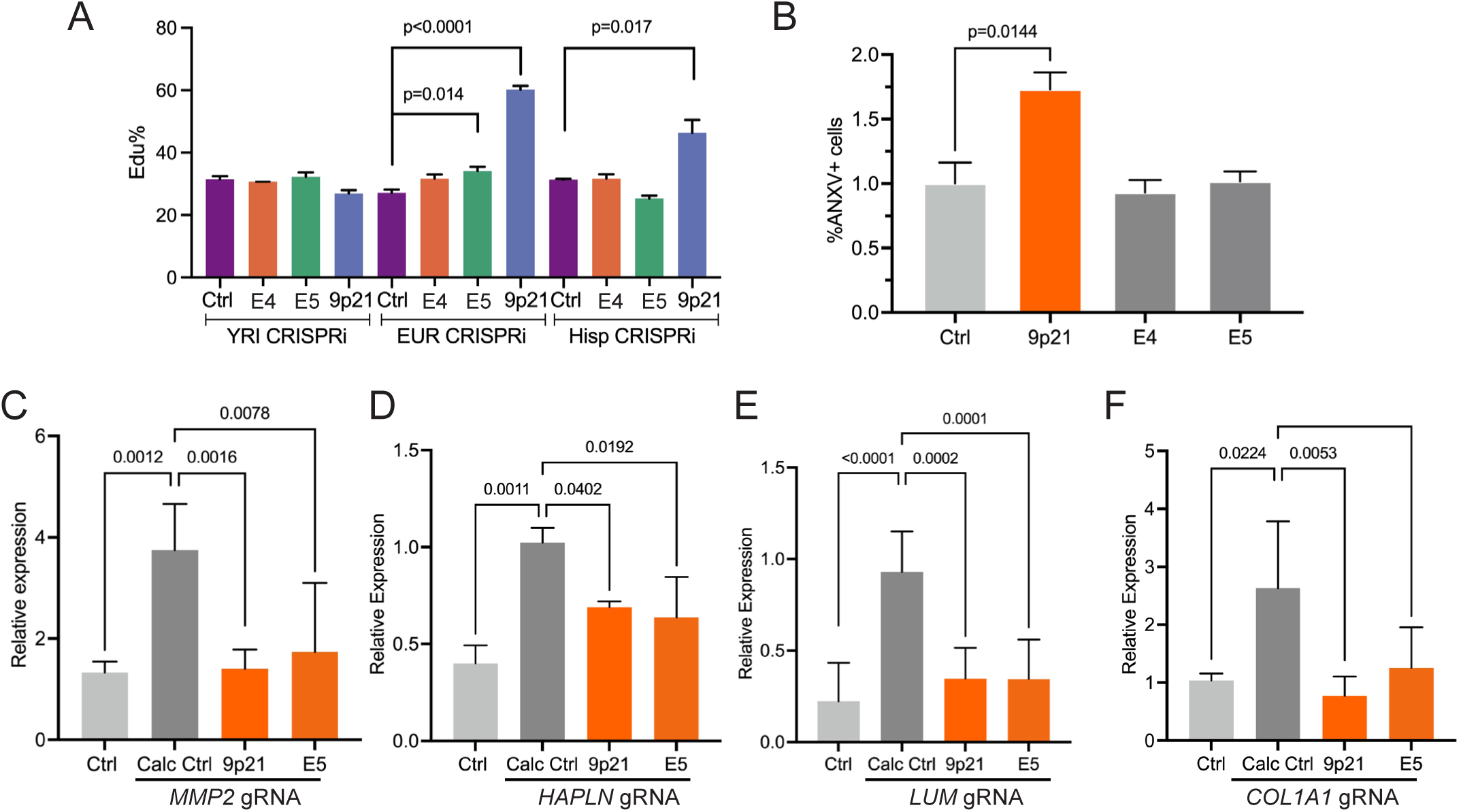
9p21.3 enhancers regulate SMC proliferation, apoptosis and vascular calcification. **A)** Lentivirus transduced gRNAs targeting individual or combined enhancers in CRISPRi-HCASMC were evaluated for proliferation with a FACS based EdU assay. EUR HCASMC heterozygous for both rs1537372 and rs4977574 showed increased proliferation in response to transduction with an individual gRNA targeting E5, as well as the combined lentiviral transduction of gRNAs targeting all 10 identified enhancers. Identical studies with EUR (Hispanic) cells with identical heterozygous genotypes also showed increased cell division with the combined gRNAs. Interestingly, an HCASMC line homozygous for the YRI alleles did not show significant changes in the level of proliferation. **B)** Lentivirus transduced gRNAs targeting individual or combined enhancers in EUR HCASMC expressing dCAS9-VPR CRISPRa (CRISPRa-HCASMC), were evaluated for apoptosis with a FACS based annexin V assay after 48 hrs. These studies revealed an increase in apoptosis in cells receiving the gRNAs targeting all combined enhancers at 9p21.3. **C)** Vascular calcification of CRISPRi-HCASMC was conducted with an in vitro calcification assay widely used to investigate the role of SMC toward plaque calcification (Methods). Well validated markers of the chondromyocyte SMC phenotype were investigated in cells maintained in basal culture medium, basal plus calcification medium, and cells transduced with the individual E5 enhancer gRNA encoding lentivirus or with lentivirus expressing gRNAs for targeting all 10 of the enhancers.

Another cell state change that has been linked to 9p21.3, cellular apoptosis ^26^, was similarly investigated with CRISPRi and CRISPRa targeting with gRNAs against individual E4 or E5 enhancers, or all enhancer combined. HCASMC were EUR with genotypes AA at rs4977574 and GG at rs1537372, transduced with enhancer targeting or negative control expressing gRNA. The assay for these studies was an annexin V-FACS based readout to quantify the number of apoptosing cells under the different conditions **(Fig. 5B)**. There was not a significant change in apoptosis for cells expressing the CRISPRi construct with either individual or combined gRNAs, so we employed CRISPRa to model enhancer activating conditions. Even with CRISPRa there was no change in apoptosis readout for the individual E4 or E5 enhancers, but there was a significant increase in apoptosis when gRNAs targeting all 10 of the enhancers identified at 9p21.3 were employed. These data suggest that this aspect of 9p21.3 function is distributed across multiple enhancers affecting expression of multiple genes at this locus.

Risk for another cardiovascular trait that maps to the 9p21.3 locus is coronary artery calcification (CAC), a surrogate marker that is widely regarded as the most sensitive clinical biomarker for CAD complications such as myocardial infarction ^37,74–76^. To investigate the relationship of this surrogate CAD marker with the causal enhancers and genes we have identified at 9p21.3, we used a well validated in vitro model of SMC vascular calcification to study the effect of knockdown of locus enhancers on gene expression for a number of genes that have been identified as markers of the chondromyocyte lineage ^77^. Enhancer CRISPRi knockdown of E5, and the combined effect of knocking down all 10 of the enhancers showed significantly decreased expression of many CMC markers *COL4A1*, *LUMICAN, MMP2* **(Fig. 5C-E)** ^8,77,78^.

## Discussion

Investigation of the mechanisms by which CAD genes residing in GWAS loci contribute to disease pathways has been slow, do to the difficulty linking disease associated variation to enhancers and risk genes that are regulated by these enhancers, i.e., V2E2G linking. To promote V2E2G discovery and promote progress in this field, we have employed multi-locus CRISPR-based enhancer Perturb-seq to identify causal relationships in CAD loci. We employed the DC-TAP-seq method to focus gene discovery efforts and study high likelihood genes in associated loci, identifying 76 enhancers and 59 genes. A number of these genes have already been the subject of mechanistic studies and serve to validate these findings, e.g., *MYO9B* ^2^, *FURIN* ^23^, *ADAMTS7* ^24^, *CDKN2B* ^25,26^*, COL4A1* ^27^, and *SMAD3* ^9^, and an additional 12 genes were validated with focused CRISPRi studies, providing an overall confirmation of ∼25% of our DC-TAP-seq findings. Gene ontology and protein-protein interaction network analyses pointed to enrichment of the TGFB pathway, as identified in previous reviews ^79^, as a dominant regulator of disease risk, involving *FURIN, LOXL1, CDKN2B, HTRA1, SMAD3*, and *STAT3* ^26,80,81^. Other pathways that showed enrichment included cartilage development, cellular proliferation, and matrix organization processes, which have been previously identified through the study of individual CAD GWAS genes that are known to function in SMC undergoing transitions to the fibromyocyte and chondromyocyte phenotypes ^8,10,11,56^. Importantly, other fundamental pathways were identified, and noted to regulate cellular functions that have been linked to smooth muscle cell and atherosclerosis disease processes, including ubiquitination ^82,83^, chaperone assisted protein folding ^84–86^, microtubule and mitotic spindle formation that is critical for the cell cycle ^87–89^.

We acknowledge that enhancers can modulate expression of more than one gene, and while our DC-TAP-seq studies have provided experimental evidence that CAD associated enhancers primarily regulate one candidate gene, the context for many of these genes supports the likelihood that they are causal toward CAD risk. Based on the inclusion criteria for our study, we know that the identified genes are expressed in SMC, and many of the genes reside in pathways and gene networks that are critical for this cellular lineage function in disease. For instance, relating to TGFB signaling, Furin serves to process and thus activate the *TGFB1* gene product, another CAD associated gene ^2,3,90^, that promotes ECM components that likely provide a stabilizing effect on atherosclerotic plaque. *LOXL1* is a paralog of the CAD candidate gene *LOX,* and both of these genes encode the CAD causal factor BMP1 processed extracellular enzymes necessary for the formation of covalent cross-links that provide collagen and elastic fibers that promote tensile strength and disease plaque stability ^91^. Another DC-TAP-seq gene *LOXL1-AS1* is an antisense lncRNA that likely regulates expression of *LOXL1* in SMC and is the target of a different enhancer than its protein coding partner. Importantly, *BMPR2* is nominated here as a causal gene for CAD, linking the known SMC hypocontractile and hyperproliferative phenotype of pulmonary hypertension, to the causality of this cell type in CAD ^92^.

*CDKN2B* was a nominated causal CAD identified locus with the multi-locus screen. This gene resides in the ∼100,000 base pair region of the genome at 9p21.3 that contributes the greatest amount of disease risk, but the mechanisms of disease risk and risk for related clinical features at this locus have not been elucidated. By conducting a focused enhancer DC-TAP-seq screen restricted to 9p21.3, we have identified a variant rs1537372 with a minor G allele that contributes to an aryl hydrocarbon receptor (AHR) binding site in the YRI genome **(Fig. 4A-D, I, Suppl. Figs. 5B)**. This allele is 10-times more frequent in the YRI ancestry genome than the EUR genome (EUR 0.44, YRI 0.04) and is not in linkage disequilibrium with any other alleles in the YRI haplotype as represented in 1000 Genomes and TopMed. We have shown that this allele contributes to AHR binding at enhancer E4, and that transcriptional response is dependent on the presence of obligate heterodimer aryl hydrocarbon receptor nuclear translocator (ARNT), and confers transcriptional response to cellular stimulation with dioxin (TCDD). These data, along with DC-TAP-seq findings that E4 regulates transcription of nearby *CDKN2B*, thus provides evidence that this V2E2G connection contributes to the described lack of CAD risk for YRI ancestry individuals at this locus. EUR and other ancestry groups have this allele at low frequency and the protective effect afforded by AHR binding, which has been shown to retard atherosclerosis development in mouse models ^56^. While the AHR locus has not been associated with disease risk, AHR has been shown to be a potent biomarker for human CAD risk ^93^, and its critical downstream regulator TIPARP resides in a CAD GWAS locus ^2,3^.

Through sex-restricted GWAS analyses, CAD risk at 9p21.3 has been shown to be largely contributed by the biological male genotype, and allelic variation identified as highly linked to this observation ^36^. At enhancer E4 we noted that rs4977574 was predicted to perturb a motif for binding of hormone receptors, most consistent with androgen receptor (AR) binding **(Fig. 4E-I, Suppl. Figs. 6A, B)**. This enhancer was linked to expression of *CDKN2B*, and this A variant specifically shown to promote AR binding and regulation of transcription in an allele-specific manner. Further, this motif was shown to confer allele-specific transcriptional response to dihydrotestosterone (DHT) stimulation. Risk at rs4977574 has not been associated with sex differences in genotype, but as these studies show, contributes to a sex-specific CAD mechanism through physiological hormonal effects that are based on biological sex.

The most significant genetic association for the CAD surrogate marker of coronary artery calcification (CAC) is for variation at 9p21.3 ^37^. While this vascular syndrome has been primarily studied in EUR populations, it is well established that individuals of African ancestry have a lower incidence of this risk factor ^94,95^. In addition, men develop CAC at an earlier age and have overall considerably more calcification than women as measured by CT scan and CAC score ^38,96^. Recent GWAS have mapped CAC genetic loci, and shown that there is considerable overlap of variation associated with both CAC and CAD, including at the 9p21.3 locus ^37^. These ancestry- and sex-related differences and relationship to risk for CAC to CAD, raise the question whether CAD variants and enhancers studied here are causal for both CAC and CAD risk.

The 9p21.3 locus has been associated with risk for vascular calcification, largely restricted to males and EUR ancestry individuals, suggesting a mechanistic relationship to the AHR and AR effects described at rs1537372 and rs4977574. Indeed, these CAD causal SNPs are also associated with CAC themselves and are in high LD with SNPs primarily GWAS associated with CAC, and may in fact be causal for this surrogate marker as well as risk for CAD. AHR has been investigated for its effects on the cell state transitions in SMC, and shown to inhibit their transition to the chondromyocyte phenotype that contributes to vascular calcification in mouse hypercholesterolemic atherosclerosis ^56^. By contrast, androgen signaling in vascular cells in the setting of mouse vascular disease models has been found to promote vascular calcification, despite inhibiting disease progression ^97^. Proof of the relationships between the AHR and AR mediated CAD risk and vascular calcification mechanisms in SMC will require further studies but promises to provide compelling data regarding this risk factor and disease.

We note that Harismendy et al. explored the role of 9p21 DNA variants associated with CAD in endothelial cells and how these variants impact interferon-γ signalling in that cell type ^98^. The study utilized human vascular endothelial cells (HUVEC) to examine the cellular mechanisms underlying this association. They identified a number of enhancers in the 9p21 region, and in HUVEC they highlighted the role of CAD risk alleles disrupting a STAT1 binding site at their enhancer 9 (ECAD9). The disruption was shown to affect the binding of STAT1, which in turn altered the expression of nearby genes, notably *CDKN2A* and *CDKN2B*. We therefore compared the coordinates of our SMC enhancers to their EC enhancers, and our enhancers 9 and 10 match with their ECAD9, with similar results of coordinating *CDKN2A, CDKN2B* and *MTAP* expression **(Suppl. Fig. 3C).** Importantly, other regions critical for SMC mechanisms of disease were not identified in their studies. These differing results suggest that different cell lineages cam employ different, as well as the same, enhancers to regulate gene expression and thus critical cell state processes that determine disease risk.

This study advances our understanding of CAD pathogenesis by identifying novel multi-locus enhancer-gene pairs and specifically at the 9p21 locus in SMCs and elucidating variant-specific mechanisms underlying disease risk. By integrating functional genomics with variant-level analyses, we highlight pathways driving vascular calcification and genetic disease risk and propose potential targets for therapeutic intervention. Our work represents a significant step toward decoding the genetic basis of CAD and its variation across sexes and ancestries.

## Methods

### Prioritization of CAD GWAS variants

Summary statistics for CAD GWAS were downloaded from the GWAS Catalog ^99^ with accession IDs GCST90132314, GCST90132315, and GCST005195. Summary statistics for the Million Veterans Program CAD GWAS were obtained from MVP investigators ^3^. The variant calls on the autosomal chromosomes 1 to 22 from the 1000 Genomes Project ^58^ samples sequenced at 30x coverage by the New York Genome Center were downloaded from http://ftp.1000genomes.ebi.ac.uk/vol1/ftp/data_collections/1000G_2504_high_coverage/working, converted to PLINK format ^100^, and filtered to those with a minor allele count of at least one in the 633 donors from European ancestry. All genome-wide significant CAD GWAS variants (p-value < 5e-8), and all 1000 Genomes Project variants from European ancestry with an LD ≥0.8 with any of the genome-wide significant CAD GWAS variants were retained for downstream analyses.

### Enhancer targeting library design and DC-TAP-seq target gene panel for multi-locus screen

For the library design, we utilized ChromBPNet ^48^ to prioritize CAD GWAS variants and variants in linkage disequilibrium (LD) with an r² ≥ 0.8 resulting in a library with 188 genetic variants. These reside in 180 distinct regulatory peaks. The variants were selected based on criteria from human and mouse in vivo SMC datasets ^5,77^, as well as human coronary artery smooth muscle cells (HCASMC). Variants were required to meet the following statistical thresholds for the ChromBPNet-derived scores in at least one of the datasets: an absolute log fold change (logFC) empirical p-value < 0.01, a Jensen-Shannon Distance (JSD) empirical p-value < 0.01, or an absolute quantile difference empirical p-value <0.01. Additionally, variants were filtered to overlap with an ATAC-seq peak identified in the 2105 telo-HCASMC. Variants located in coding or splicing regions, as well as those situated within promoter regions, were excluded from the library. Furthermore, to ensure potential regulatory relevance to HCASMC, each variant had to be located within 100 kb of the transcription start site (TSS) of at least one gene expressed in HCASMC or be linked to such a TSS via Hi-C data or be located within 250kb of at least one curated gene expressed in HCASMC.

In addition to the targeted variants, we designed negative controls for the targeted loci, where each negative control consisted of a 500 bp genomic region located between 2 kb and 10 kb away from the associated regulatory peak. These regions could not intersect any annotated regulatory peaks or exons. If the selected regulatory peak was in an intronic region, the negative control had to also be intronic within the same gene. Conversely, if the peak was intergenic, the negative control was required to be intergenic and could not overlap any annotated genes. Among candidate regions that satisfied these conditions, we selected the final negative control based on the lowest maximum ATAC-seq (Tn5) insertion counts across the 500 bp region. In cases where multiple candidates had similar ATAC-seq profiles, the region with the most comparable GC content to the regulatory peak was chosen to minimize sequence composition bias. This strategy yielded suitable negative controls for 176 out of the 180 selected peaks. For the remaining 4 peaks, which were too small to meet the distance and location criteria, negative controls were manually selected to maintain consistency with the overall design. For the positive controls, we used a set of regions previously identified as positive controls in the K562 screen ^21^. We further filtered these regions to include only those with at least 80% overlap with regulatory peaks identified in the 2105 HCASMC and then matched them with the loci and target genes of our selected enhancers. For each locus, we also included at least one gene transcription start site (TSS) targeted using prevalidated guide RNAs from the Weissman library^101^.

### ChromBPNet model training

We trained ChromBPNet models on pseudobulk ATAC-seq data from each mouse and human cell type, along with the data from the telo-HCASMC cell line. For training each model, we used the ATAC-seq peak and tagalign files, along with a Tn5-bias model trained on the single cell cluster from each dataset with the highest number of mapped reads. The code repository for training ChromBPNet models is at https://github.com/kundajelab/chrombpnet.

We used a 5-fold cross-validation scheme to train each model, with the chromosomes included in the train, validation, and test splits for each fold in human and mouse specified below.

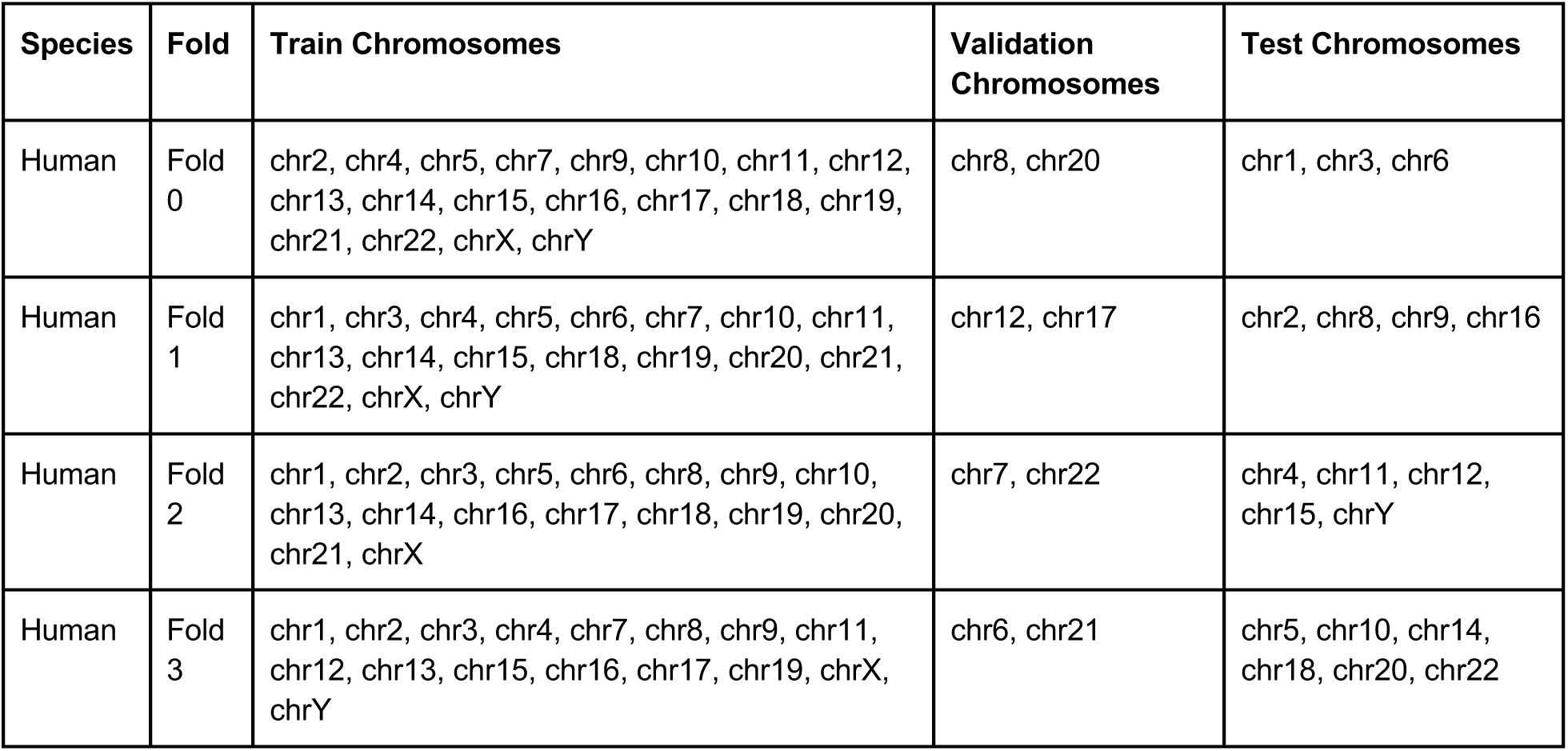
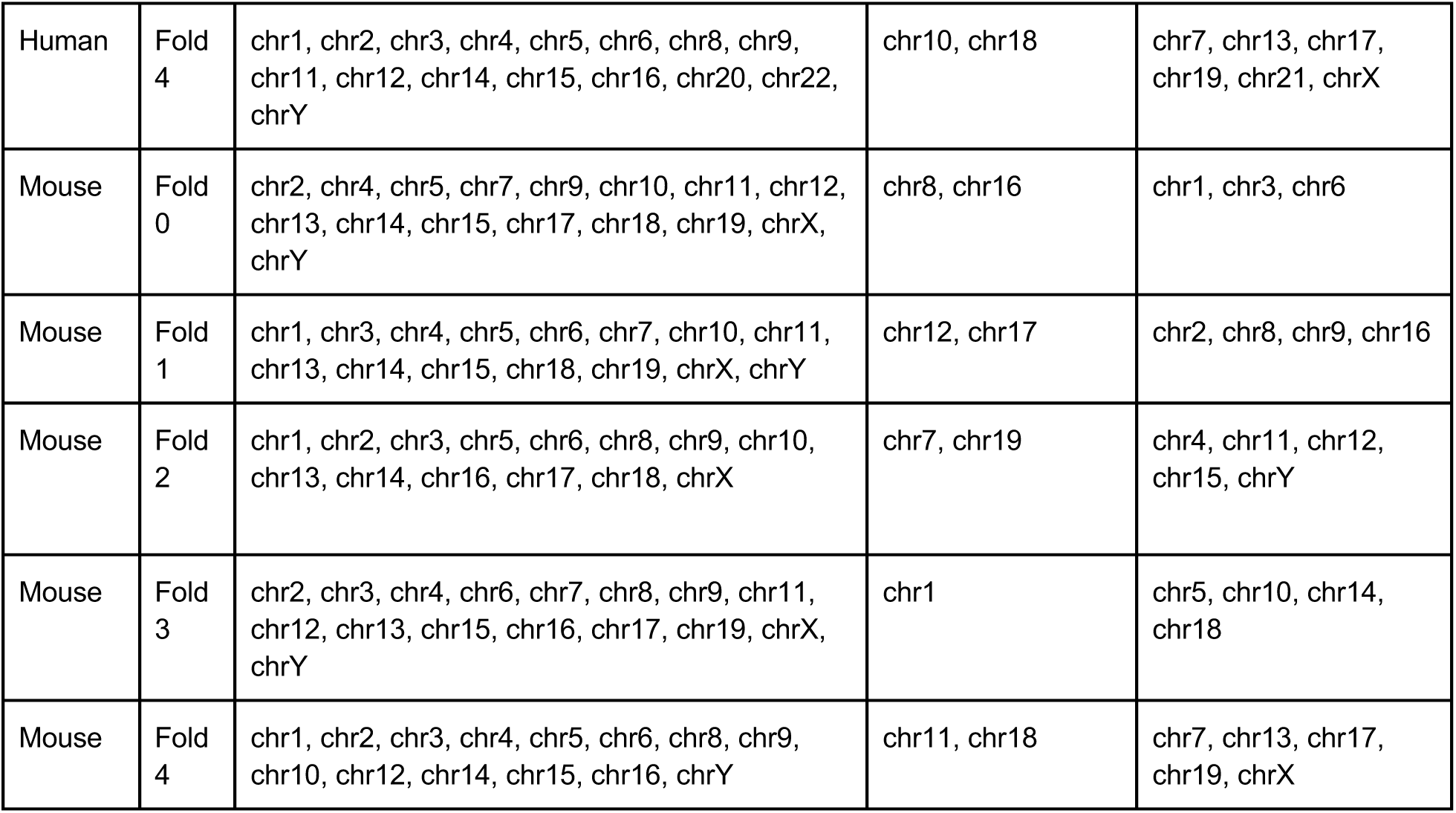

### Variant effect prediction using ChromBPNet models

We used each cell type-specific ChromBPNet model to score each CAD-associated variant by predicting the base-resolution pseudobulk ATAC-seq coverage profiles for the 1 kb DNA sequence centered at each variant and containing the reference and alternate allele. We estimated each variant’s effect size using three measures: (1) the log2 fold change in total predicted coverage (total counts) within each 1 kb window for the alternate versus reference allele, (2) the Jensen–Shannon distance (JSD) between the base-resolution predicted probability profiles for the reference and alternate allele, and (3) the absolute quantile difference, which is the difference in the quantile of the predicted total counts between the alleles with respect to the distribution of the predicted total counts for all peaks from the cell type where the model was trained.

Statistical significance for each score was established using empirical null distributions constructed by shuffling the input sequence around each variant, while preserving dinucleotide frequency, re-inserting both alleles of the variant at the center of the shuffled sequence, repeating the process 1 million times to create a million null examples, and then scoring each null variant using the same methods used for the observed variants. Then, for each observed variant and for each score, we computed the proportion of null variants with an equally high or higher score to derive empirical *P*-values for the log2 fold change, JSD, and absolute quantile difference scores. The code repository for scoring variants is at https://github.com/kundajelab/variant-scorer.

### DC-TAP-seq primer panel design

Target genes for DC-TAP-seq amplification were assigned for each variant, defined as any gene located within 100 kb of the peak or two closest genes if further away than 100kb. If there were no genes within 100 kilobases the two closest genes were studied. This analysis yielded 412 enhancer target genes, which were combined with 44 control genes that do not reside in these loci and considered as “housekeeping” genes, as well as 14 validated CAD genes, resulting in 470 genes in total for the complete panel (Suppl. Tables 2-3). For the 9p21 screen, we designed DC-TAP-seq primers for all genes in the locus that had expression in telo-HCASMC.

DC-TAP-seq primers were designed using the customized pipeline described ^16^, and based on protein-coding exon annotations for the genes targeted (Gencode GRCh38.89). Available SMC 10x scRNA-seq data from the telo-HCASMC line was used to identify expressed isoforms and likely polyadenylation (polyA) sites for every target gene of interest. If multiple polyA sites were found, the most downstream site was chosen. The most downstream annotated 3’ end was used if no polyA site could be identified. If several transcript isoforms overlapped with the inferred polyA sites or no polyA sites were found, their annotations were merged (union) to form a consensus annotation for primer design. After the automated pipeline, the annotation was also manually checked using the IGV browser.

Transcript sequences were extracted (hg38) for all target genes and nested forward primers were designed around the 3’ end: Outer forward primers were selected to result in 350-500 bp amplicons, which were then used to design inner forward primers to yield 150-300 bp amplicons. Ten forward and reverse primers were designed per target gene and aligned against human genome and transcriptome databases using blast to identify potential off-target annealing. The inner and outer primers with the fewest exonic, intronic and intergenic off-target alignments (in that order) were selected for every gene, and all primers were assessed for multiplex suitability with Primer3’s check_primers functionality. Primers were synthesized as separate oligos and purified by desalting (IDT) and combined into a common pool. DC-TAP-seq gene panel primer sequences are listed in **Suppl. Tables 3, 4**. Before the actual experiment the DC-TAP-seq panels (multi-locus and targeted 9p21-screens) were validated by amplifying a telo-HCASMC scRNAseq dataset and correlating the sequencing read counts to the transcripts per million values in telo-HCASMC bulk RNAseq.

### Guide sequence tiling and design

Guide library design was performed using CRISPRDesigner and CRISPRi tiling as described and includes ten sgRNA guides tiled around the summit of the enhancer peak ^21^. The library consisted of 5299 sgRNAs, of which 20% were negative guides and non-targeting guides. (**Suppl. Table 1**). For the 9p21 screen, the library consisted of 200 guides of which 10 were non-targeting guides, 20 were negative guides and 10 guides per TSS site targeted (**Suppl. Table 6**).

### Cloning of the guide library

The 3’ direct capture Perturb-seq plasmid (pBA904, Addgene #122238) was digested with BstXI and BlpI, and purified from agarose gel using QIAquick Gel Extraction Kit (QIAGEN). sgRNA DNA oligos were annealed, ligated into the digested 3’ direct-capture Perturb-seq backbone, and transformed using electroporation with Mega-X bacterial cells. Plasmid DNA was prepared and scraped from cultures plates. The libraries (multi-locus and 9p21 guide libraries) were sequenced to confirm the even distribution of sgRNA sequences in the library, with minimum and maximum number of guide copy counts falling within 2.8-fold difference.

### Lentiviral packaging

Lentiviral gene expression vectors were co-transfected with second generation lentivirus packaging plasmids, pMD2.G and pCMV-dR8.91, using Lipofectamine 3000 (Thermo Fisher, L3000015) according to the manufacturer’s instructions. ViralBoost Reagent (AllStem Cell Advancements, VB100) was added (1:500) with fresh media after 5 h. Supernatant containing viral particles was collected 48 h after transfection and filtered.

### Cell culture

Primary HCASMC were purchased from Cell Applications, Inc (San Diego, CA) and Lonza BioScience and were cultured in complete smooth muscle basal media (Lonza, #CC-3182) according to the manufacturer’s instructions. All experiments with primary HCASMC were performed between passages 5–8 with cell lots 1386, 9071501, 2989, 1508, 2105 and 2305. Rat aortic smooth muscle cells (A7r5) and human embryonic kidney 293 cells (HEK293) were purchased from ATCC and cultured in Dulbecco’s Modified Eagle Medium (DMEM) high glucose (Fisher Scientific, #MT10013CV) with 10% FBS at 37 °C and 5% CO2. A7r5 at passage 6-18 were used for experiments.

### telo-HCASMC with dCas9-KRAB-ZIM3 or dCas9-VPR

hTert immortalized human coronary artery smooth muscle cells (telo-HCASMC) obtained from the Clint Miller lab were utilized for in vitro experiments in this study. CRISPRi-HCASMC were generated from the telo-HCASMC by transduction at low MOI with virus encoding dCas9-KRAB-ZIM3 (*pHR-UCOE-EF1a-Zim3-dCas9-P2A-mCherry)* made from Addgene # 188766. Cells that were mCherry positive were sorted and oligoclonally expanded and maintained. The cells were transfected with the pooled sgRNA virus to obtain an MOI of 3 (titrated with ddPCR), harvested after seven days, and FACS sorted (BFP+, mCh+). For CRISPRa studies, telo-HCASMC were transduced with Addgene # 154193 (*dCas9-VPR_P2A_mCherry*), FACS sorted for mCherry positive cells and then oligoclonally expanded and maintained.

### Digital droplet PCR

We used digital droplet PCR to determine the multiplicity of infection (MOI). First, we isolated genomic DNA from telo-HCASMC. We performed droplet digital PCR (ddPCR) by amplifying both the scaffold in the direct capture vector and albumin as a reference per cell and used Bio-Rad Custom ddPCR Assay reagents. Readout was performed using the QX200 Droplet Reader and Quantasoft Software (BioRad) to determine the total number of copy number variants (CNV) of the virus in a cell.

### DC-TAP-seq library preparation using a modified 10x Genomics platform

The protocol described here was modified to be used with 10x Genomics 3’ reagent kit v4, but can be used for v3 chemistry as well. Library preparation follows 10x protocol for GEM-RT incubation. Cell input was set to a targeted cell recovery of 20,000 cells per lane. Cell counts were double-checked directly before loading on the 10X Genomics Chromium controller. After GEM-RT incubation, cDNA was cleaned up according to the 10x protocol and the eluent used as input for DC-TAP-seq protocol PCR1 (outer primers), followed by inner primer PCR2 step. PCR3 step added Illumina adapters to the product to enable sequencing of the library which was double indexed (i5 and i7). The DC-TAP-seq protocol produced Illumina sequence ready libraries identical to 10x Genomics single-cell 3’ libraries using standard Illumina Read 1 and Read 2 primers. The detailed protocol of all the steps is deposited in Protocols.io: (https://www.protocols.io/blind/9BFC6CED8F1211EF8F770A58A9FEAC02).

### SCEPTRE analysis

The DC-TAP-seq FASTQ files underwent processing using Cell Ranger version 8.0.0. A custom transcriptome incorporating the DC-TAP-seq targeted gene panel was generated with Cell Ranger mkref. For Cell Ranger count run, a feature reference CSV file specified the gRNA sequences and cloning construct. Subsequently, the processed gene expression and gRNA count matrices were imported into sceptre v0.10.0.

In the power test, gRNAs targeting the TSS region or enhancers linked to known genes served as positive controls, while non-targeting and safe-targeting control gRNAs served as negative controls during calibration. Discovery of enhancer-to-gene pairs was restricted to pairs within 1M bp distance. Covariate analyses considered gRNA UMI counts, cell numbers per gRNA sequence, gene expression UMI counts, cell numbers per detected gene, library barcode, and Seurat clusters. Mixture modelling ^102^ was employed for gRNA assignment. When multiple gRNA sequences targeted the same enhancer, all gRNAs were aggregated for testing. Permutation tests were conducted for enhancer-gene discovery, with statistical tests corrected using Benjamini-Hochberg FDR=10%. The intersection of DC-TAP-seq and scE2G predicted enhancer-gene connections was determined by requiring enhancers to overlap by at least 1bp and be linked to the same genes.

### Power calculations

Power analyses were performed using the simulation based approach from Ray et al., 2025:https://github.com/EngreitzLab/DC_TAP_Paper/tree/main. Briefly, to parameterize our simulations, we modeled gene expression using a negative binomial distribution, with dispersion estimates calculated by SCEPTRE and baseline mean expression and size factors calculated using DESeq2. Synthetic UMI counts were then drawn from the negative binomial distribution where the mean expression for each gene in perturbed cells was shifted by a defined effect size that incorporated centered, guide-level variability sampled from a normal distribution (σ=0.13). UMI counts for unperturbed control cells were drawn from the same Negative Binomial distribution without changing the mean. The standard SCEPTRE discovery analysis was subsequently run on 100 iterations for each effect size and high-quality element-gene pair, and we reported the fraction of successful discoveries with negative effects as the power for that pair.

### Benchmarking analyses large-scale screen

The use of precision-recall and PoPs analyses were previously used to assess Perturb-seq analyses ^103^. PoPS is a method to nominate probable causal genes in a GWAS locus and we applied PoPS to previously published summary statistics using a predefined group of gene sets as previously described ^2^. For each GWAS signal, we calculated the PoPS rank among ‘nearby genes’ (2 to either side of the lead SNP, and all within ±250 kb). To evaluate precision recall, eight gold standard SMC CAD genes (*CDKN2B, TCF21, SMAD3, BMP1, LOXL1, MYO9B, PALLD, FN1*) were used to benchmark the performance of TAPseq or other studies in identifying known causal genes. Precision was defined as the number of gold standard genes identified divided by the total number of genes called within the corresponding loci. Recall was defined as the proportion of gold standard genes successfully identified in the study ^103^.

### Differential expression analysis using MAST

Sequencing reads were down-sampled without replacement to a defined average genic read depth per cell. Then, we sampled 10 to 150 cells expressing each gRNA and performed DE against 500 cells randomly sampled from the scrambled gRNA cells, repeating the process 100 times. Precision-recall curves were computed across all sampling runs with a defined read and cell number, by assuming that the intended gRNA target is the only true positive. MAST ^104^ with the number of genes observed per cell as a covariate was used as a DE test.

### Gene ontology analysis

To identify biological processes, molecular functions, and cellular components enriched in our dataset, we performed Gene Ontology (GO) enrichment analysis using the genes identified in the DC-TAP-seq multi-locus screen. Significant GO terms were ranked by p-value and fold-enrichment, and key biological processes were highlighted based on relevance to the study objectives. TF motif analysis was conducted using HOMER findMotifGenome.pl. The 300-bp regions from the enhancers of significant E2G pairs were extracted and analyzed. Non-significant enhancer regions served as the background for the motif analysis. The JASPAR 2020 database was used as the reference, and the known motifs results were used for presentation.

### HCASMC phenotypic proliferation, apoptosis and calcification assays

For the proliferation assay, EdU was introduced into telo-HCASMC culture 3 hours before assay for uptake. The protocol for Click-iT Plus EdU proliferation kit (Thermo Fisher) was followed as instructed. For the apoptosis assay, telo-HCASMC were treated with Doxorubicin (1uM) for 24 hours to induce apoptosis. The RealTime-Glo Annexin V Apoptosis kit (Promega) was used as instructed to quantify the degree of apoptosis in telo-HCASMC. To assay for calcification, these cells were exposed to calcification media with 10mM beta-glycerophophate and 100ug/ml ascorbic acid as described previously ^56^.

### Single guide validation

The most effective gRNA sequence targeting an enhancer was individually cloned into the pBA904 vector and transfected into CRISPRi-HCASMC. After five days, RNA was collected, extracted, and converted to cDNA. The effect of enhancer targeting was then assessed by comparing the expression levels of the target genes *CKB, PPP1R18, RAB23, EDEM2, MICA, TP53INP2*, (**Fig. 1 I-K, and Suppl. Fig. 2D-F),** as well as *LOXL1, MYO9B, NAV1, FURIN, SBF2,* and *PALLD* (data not shown) to the control.

RNA was isolated using RNeasy plus micro kit (Qiagen, #74034) and total cDNA was prepared using High-capacity RNA-to-cDNA kit (Life Technologies, #4388950). Gene expression was assessed using TaqMan qPCR probes (Thermo Fisher) for *CDKN2A* (HS99999189_M1), *CDKN2B* (Hs00793225_m1), *CDKN2B-AS1* (Hs03300540_m1), *MTAP* (HS00559618_M1), MICA (Hs07292198_gH), SBF2 (Hs00959709_g1), *TP53INP3* (Hs00894008_g1), *RAB23* (Hs00212407_m1), *PPP1R18* (Hs00292978_m1), *CKB* (HS00176484_M1) according to the manufacturer’s instructions on a ViiA7 Real-Time PCR system (Applied Biosystems, Foster City, CA). Relative expression was normalized to *UBC* (Hs00824723_m1) levels.

### CHIP experiments

All experiments were performed on primary HCASMC between passages 4 and 7. Antibodies used for ChIP-quantitative PCR (qPCR) were all pre-validated according to ChIPseq guidelines. Purified mouse monoclonal antibody against human Ahr (SantaCruz-133088) and rabbit polyclonal AR (EpiCypher 13-2020) antibody were used. Cell lines with various genotypes of rs4977574 and rs1537372 were used for haplo-ChIP. Approximately 2,000,000 HCASMC for each line were fixed with 1% formaldehyde and quenched by glycine. The cells were washed three times with PBS and then harvested in ChIP lysis buffer (50 mM Tris-HCl, pH 8, 5 mM EDTA, 0.5% SDS). Crosslinked chromatin was sheared for 3 × 1 min by sonication (Branson SFX250 Sonifier) before extensive centrifugation. Four volumes of ChIP dilution buffer (20 mM Tris-HCl, pH 8.0, 150 mM NaCl, 2 mM EDTA, 1% Triton X-100) was added to the supernatant. The resulting lysate was then incubated with Dynabeads™ Protein G (Thermo Scientific, 10009D) and antibodies at 4 °C overnight. Beads were washed once with buffer 1 (20 mM Tris pH 8, 2 mM EDTA, 150 mM NaCl, 1% Triton X100, 0.1% SDS), once with buffer 2 (10 mM Tris pH 8, 1 mM EDTA, 500 mM NaCl, 1% Triton X100, 0.1% SDS), once with buffer 3 (10 mM Tris pH 8, 1 mM EDTA, 250 mM LiCl, 1% NP40, 1% sodium deoxycholate monohydrate), and twice with TE buffer. DNA was eluted by ChIP elution buffer (0.1 M NaHCO_3_, 1% SDS, 20 μg/ml proteinase K). The elution was incubated at 65 °C overnight, and DNA was extracted with a DNA purification kit (Zymo D4013). The purified DNA from various lines were analyzed by quantitative PCR using the ABI ViiA 7, with the same primer applied to each SNP. Assays were repeated at least three times. Data shown were average values ± SD of representative experiments. Primer sequences are listed in **Suppl. Table 10**.

### Dual luciferase assays

Genomic regions, as listed in **Suppl. Table 11,** were synthesized by Twist Biosciences and then cloned into pLuc-MCS vector. Human embryonic kidney (HEK) or A7r5 rat aortic SMC were seeded into 24 well plate (2×10^4^ cells/well) in DMEM containing 10% FBS and incubated at 37 °C and 5% CO_2_ overnight. Cells were transfected with luciferase reporter plasmids, transcription factors and Renilla plasmid using Lipofectamine 3000 (Invitrogen, #L3000015). Five hours after transfection, the media was changed to fresh complete media. Relative luciferase activity (firefly/*Renilla* luciferase ratio) was measured using Dual Luciferase Reporter Assay System (Promega, #E1960) on a SpectraMax iD3 luminometer (Molecular Devices) 24 hours after transfection. All experiments were conducted in triplicate and repeated at least 4 times. Dioxin, DHT, estradiol or progesterone were added at the time of transfection and maintained until luciferase activity was measured. Their respective concentrations are given in the figure legends.

### Statistics & Reproducibility

All statistical analyses were conducted using GraphPad Prism software version 9. Difference between two groups were determined using an unpaired two-tailed *Student’s t-test.* Differences between multiple groups were evaluated by one-way analysis of variance (ANOVA) followed by Dunnett’s post-hoc test after the sample distribution was tested for normality. *P* values <0.05 were considered statistically significant.

## Data Availability

Data described in this manuscript have been uploaded to the Data Coordinating Center of the Impact of Genomic Variation on Gene function, and will be publicly available upon publication.

https://github.com/EngreitzLab/DC_TAP_Paper/tree/main

## Data availability

Data generated through these studies have been uploaded to IGVF portal/PATH and will become publicly available coincident with publication of this manuscript. Source data are provided with this paper, in the Source Data Files.

## Acknowledgements

This work was supported by National Institutes of Health grants R01 HL171045 (TQ), R01HL134817 (TQ), R01HL139478 (TQ), R01HL156846 (TQ), R01HL158525 (TQ), UM1 HG011972 (TQ+JME), and the William G. Irwin Foundation (TQ), as well as a Human Cell Atlas grant (ZF2019-002437) from the Chan Zuckerberg Foundation (TQ). M.R. received support from the Sigrid Juselius Foundation and the Emil Aaltonen Foundation. This work was also supported by American Heart Association grant 24POST1193881/MarkusRamste/2024. E.J. and J.R. were supported by the Novo Nordisk Foundation Center for Genomic Mechanisms of Disease (NNF21SA0072102). J.M.E. was also supported by NHLBI R01HL159176; the Applebaum Foundation; and the BASE Research Initiative at the Lucile Packard Children’s Hospital at Stanford University. M.U.K. was supported by the European Union (ERC, SECRET, 101125115). Views and opinions expressed are however those of the author(s) only and do not necessarily reflect those of the European Union or the European Research Council. Neither the European Union nor the granting authority can be held responsible for them.

## Author contributions

MR performed experiments and wrote the initial manuscript version.

CW helped construct the 9p21 screen, and collaborated on manuscript preparation.

SK performed numerous analyses, including the ChromBPNet work.

QZ performed analyses of SCEPTRE and MAST data outputs, and helped with analyses of genomic data.

DL provided extensive mouse single cell RNA sequencing data for neural network analyses, and provided guidance regarding integration of disease single cell RNA sequencing data.

KB performed the statistical power analysis on the SMC TAP-seq and 9p21 locus discovery pairs, and produced figures describing the power analysis results.

AR helped with analyses of the multi-locus studies, and provided unpublished data regarding causal gene identification in CAD loci.

EJ provided unpublished enhancer perturb-seq pilot data, and guidance regarding Perturb-seq design.

JR provided unpublished enhancer perturb-seq pilot data, and guidance regarding Perturb-seq design.

RDC helped with figures and edited the manuscript.

GJ provided unpublished enhancer perturb-seq pilot data, and guidance regarding Perturb-seq design.

AG provided extensive support for design of the DC-TAP-seq studies, primer design, utilization of analysis pipelines for data analyses etc.

NL performed qPCR validation of CRISPR results, and performed variant editing experiments. TN performed numerous experiments with the CRISPRa-HCASMC, including validation of the 9p21 screen. She also performed luciferase reporter studies in A7R5 rat cells.

JMA provided unpublished human coronary artery multi-ome data and supported the enhancer Perturb-seq analyses.

CYP provided key input regarding the direct capture CRISPRi protocols, gRNA design, etc.

JBK provided updated Ahr knockout mouse atherosclerosis single cell data and contributed to AHR studies.

MUK provided MPRA data related to variation at 9p21.3 and edited the manuscript.

NOS provided unpublished human coronary artery multi-ome data and supported the enhancer Perturb-seq analyses.

LS provided support for the DC-TAP-seq study design.

AK supported advanced analyses with neural network tools to identify regulatory variants that modulate chromatin accessibility in CAD loci.

JME provided details for his unpublished enhancer DC-TAP-seq studies, extensive advice on the design and utilization of CRISPRi methods and guide design, and conducted statistical power analyses.

TQ conceived of these studies, edited the manuscript, created figure panels, and oversaw all aspects of the research.

## Supplemental Tables

**Suppl. Table 1.** Multi-locus CAD enhancers and targeting gRNA sequences

**Suppl. Table 2**. Multi-locus screen DC-TAP-Seq target gene list

**Suppl. Table 3.** Multi-locus screen DC-TAP-Seq PCR primers

**Suppl. Table 4.** K562 control DC-TAP-Seq PCR primers

**Suppl. Table 5.** Multi-locus screen SCEPTRE results

**Suppl. Table 6.** 9p21.3 screen enhancer coordinates and gRNA sequences

**Suppl. Table 7.** 9p21.3 DC-TAP-Seq PCR primers

**Suppl. Table 8.** 9p21.3 SCEPTRE and MAST results

**Suppl. Table 9.** 9p21.3 V2E2G links, CAD variants and MAFs across ancestries, putative disrupted TF motifs

**Suppl. Table 10.** Chip qPCR primer sequences

**Suppl. Table 11.** Genomic reporter gene sequences to measure allele-specific transcriptional activity

## Supplemental figures

**Suppl. Figure 1.**
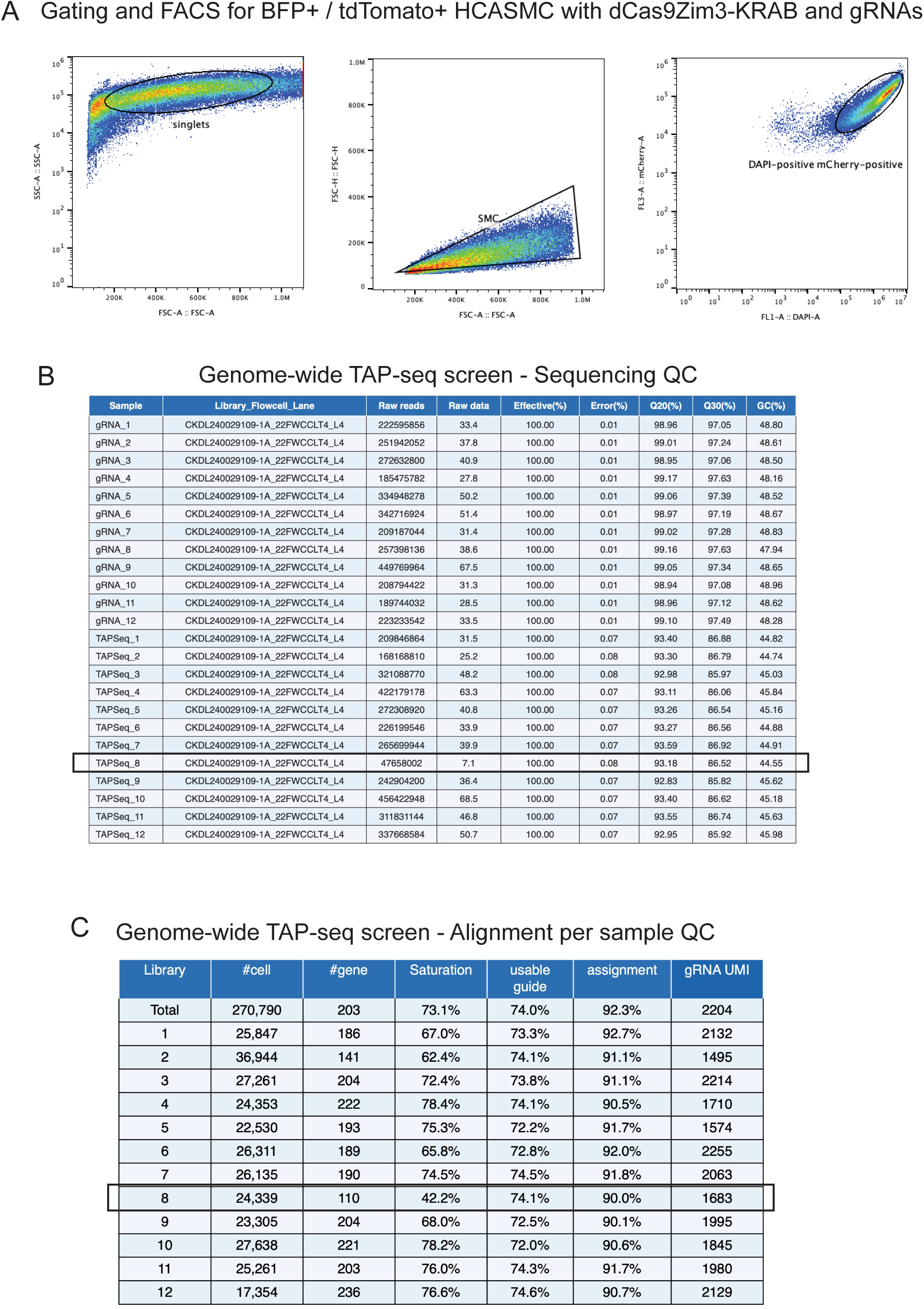
**A)** Gating and FACS for BFP+ tdTomato+ HCASMC with dCas9-KRAB-ZIM3 and gRNAs. **B)** Sequencing QC for individual captures and libraries from multi-locus DC-TAP-seq screen. **C)** Alignment of reads per library for the multi-locus DC-TAP-seq screen.

**Suppl. Figure 2.**
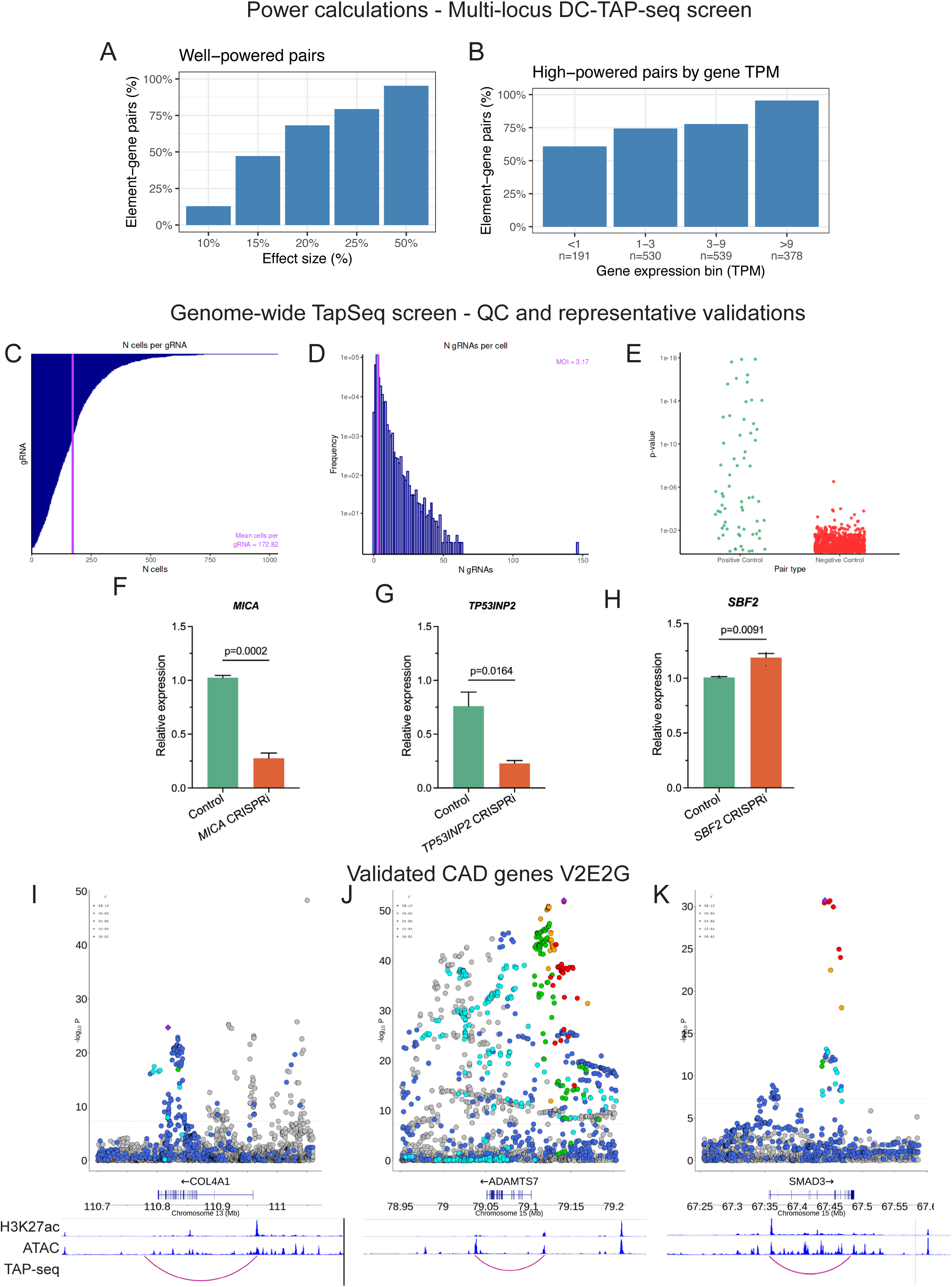
Power calculations and quality metrics of the multi-locus DC-TAP-seq screen. **A)** Percentage of all tested element-gene pairs that were well-powered, defined as having ≥ 80% statistical power, to detect different effect sizes on gene expression, in the multi-locus DC-TAP-seq experiment. 79% of pairs were well-powered to detect a 25% decrease in gene expression. **B)** Percentage of element-gene pairs that were well-powered given a 25% effect size, binned by level of gene expression in transcripts per million (TPM). The total number of element-gene pairs (n) in each bin is noted on the x-axis. **C)** Number of cells identified per gRNA. **D)** Number of gRNAs per cell. **E)** SCEPTRE calibration employed for identifying significantly differentially expressed genes, for positive and negative control cells. **F-H)** CRISPRi – PCR validation of candidate causal CAD genes identified in the multi-locus screen. **I-K)** LocusZoom plots and V2E2G links for validated GWAS CAD genes *COL4A1, ADAMTS7*, and *SMAD3*.

**Suppl. Figure 3.**
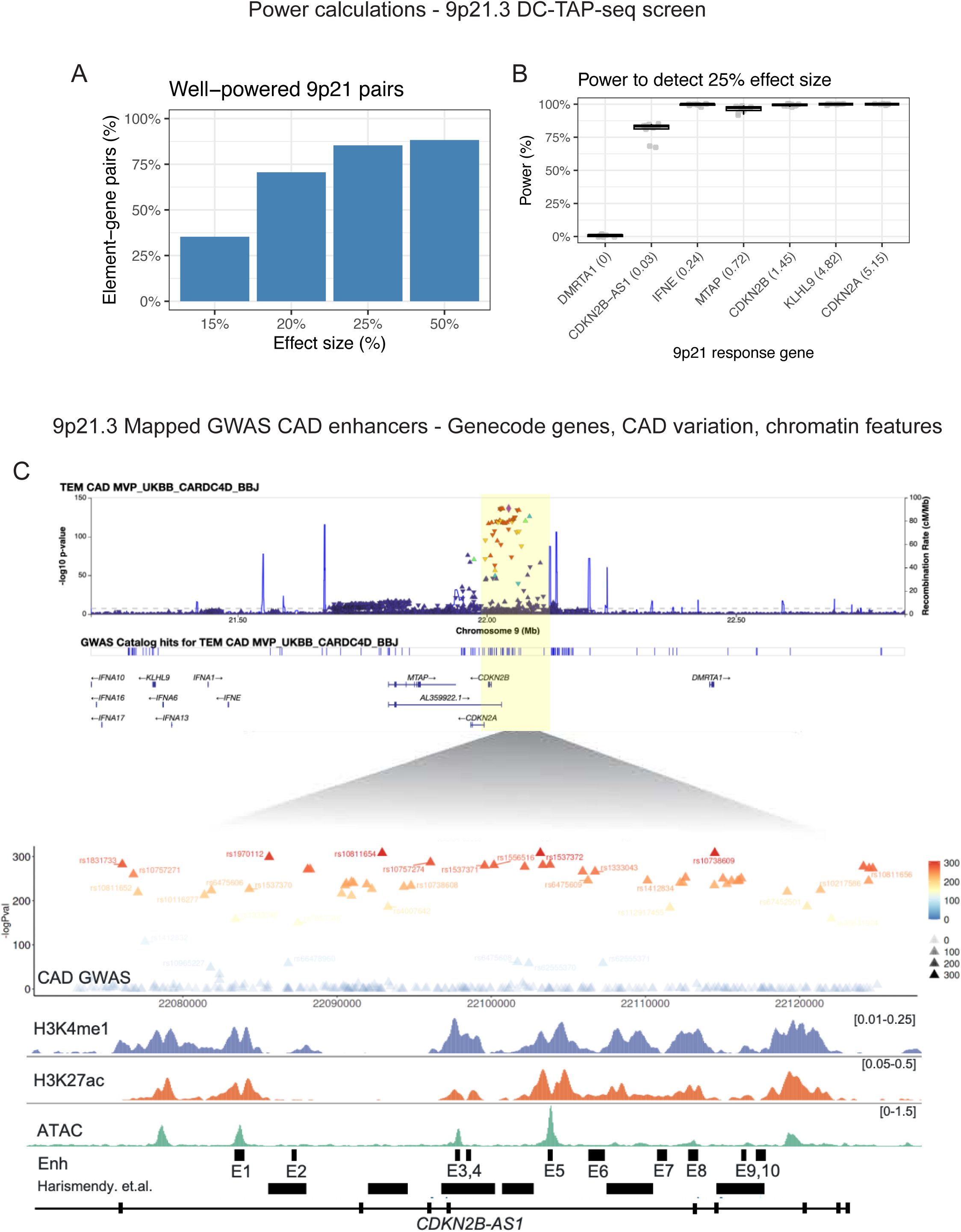
Statistical power analysis and genome browser depiction of the 9p21.3 locus architecture. **A)** Percentage of all tested element-gene pairs that were considered well-powered, defined as having ≥ 80% statistical power, to detect different effect sizes on gene expression in the 9p21 DC-TAP-seq experiment. 85% of pairs were well-powered to detect a 25% decrease in gene expression. 68 element-gene pairs were considered at the 9p21 locus (2 pairs from the set of 10 elements x 7 genes did not pass quality control). **B)** Percentage of element-gene pairs that were well-powered for each of the seven 9p21 locus genes. The two genes with lower power (*DMRTA1*, *CDKN2B-AS1*) are also very lowly expressed. The expression level of the gene (TPM) is indicated in parentheses. **C)** Genome browser depiction of the 9p21.3 locus architecture, location of causal variation and genes, CD-TAP-seq identified GWAS CAD enhancers, HCASMC histone marks and ATAC regions of open chromatin. Also shown is overlap of SMC DC-TAP-seq enhancers with those identified in endothelial cells by Harrismenday et al. are indicated.

**Suppl. Figure 4.**
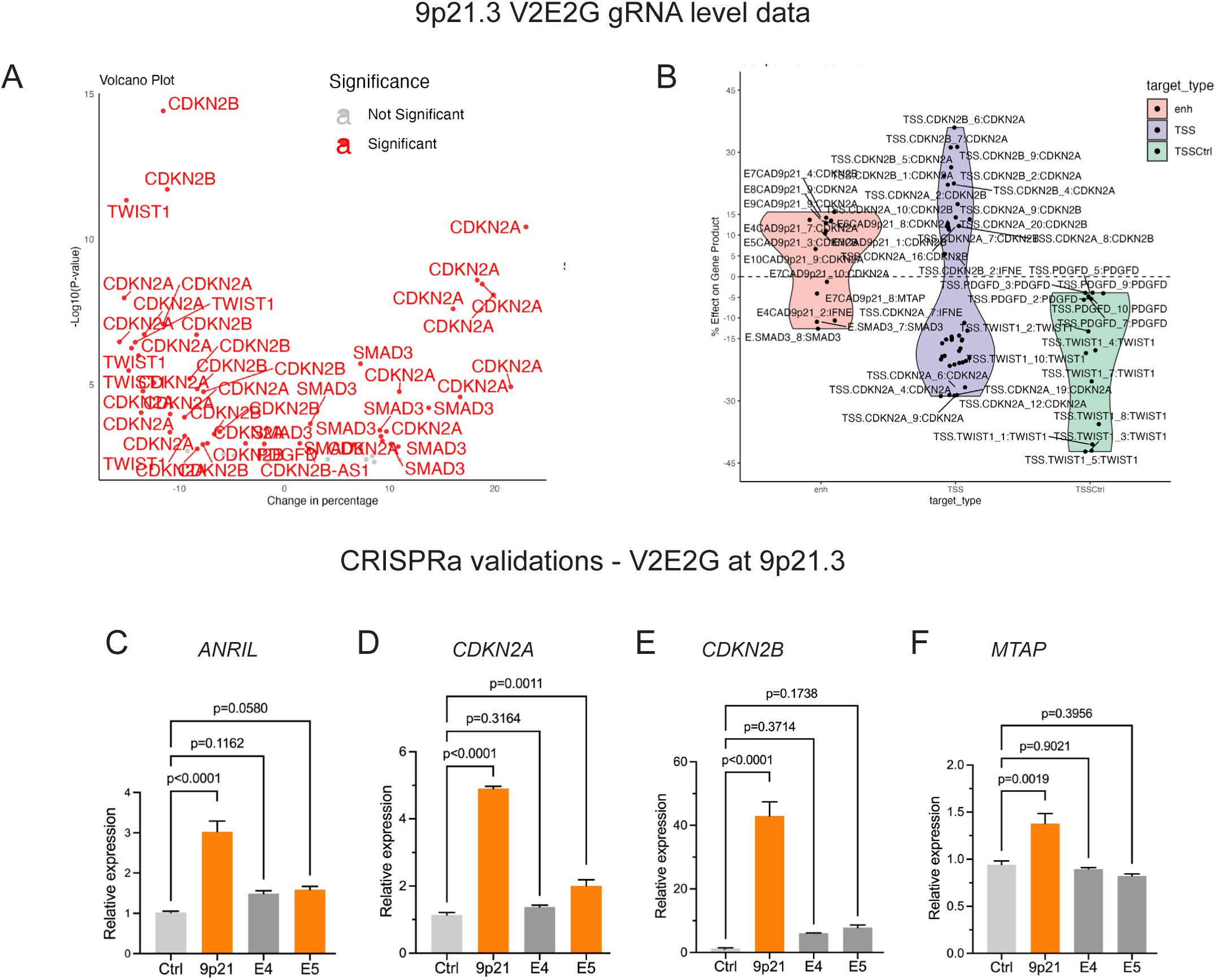
CRISPR identifications of V2E2G at 9p21.3. **A, B)** 9p21.3 V2E2G individual gRNA level DC-TAP-seq data. **C-F)** CRISPRa validations for V2E2G for *ANRIL*, *CDKN2B, CDKN2A, and MTAP*.

**Suppl. Figure 5.**
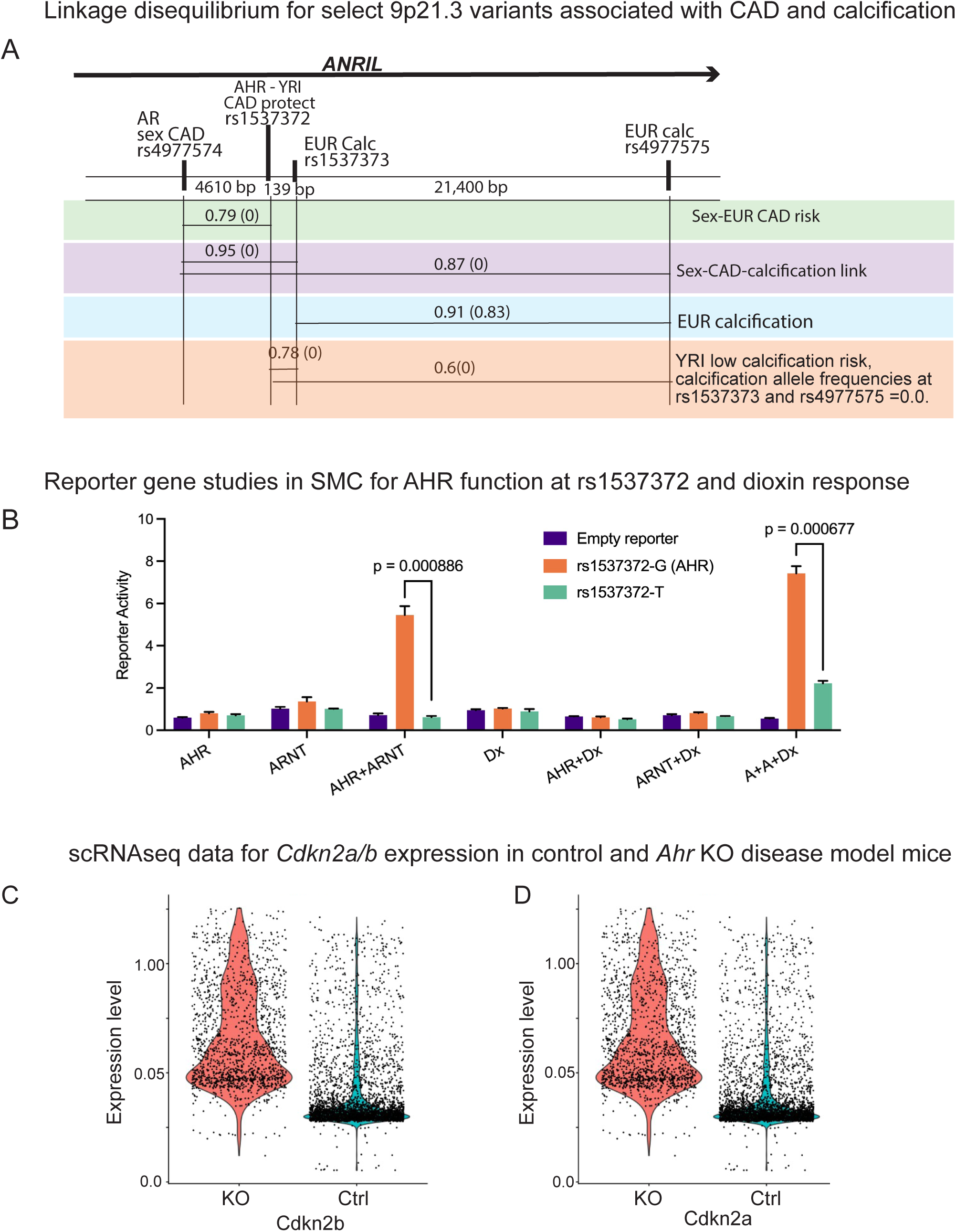
Haplotype relationships and study of transcriptional activity of AHR in cultured cells and mouse disease model. **A)** Linkage disequilibrium (LD) for select 9p21.3 variants associated with CAD and calcification. LD for EUR alleles is shown for individual pairs of SNPs that have been identified as contributing to risk for CAD and vascular calcification, with YRI LD values indicated for highlighted SNPs shown in parentheses. **B)** AHR mediated transcriptional expression linked to rs1537372 with reporter gene studies in A7R5 rat SMC. **C)** Singe cell RNA sequencing data indicates an inhibitory role of Ahr toward expression of *Cdkn2b* and *Cdkn2a* in a mouse atherosclerosis disease model.

**Suppl. Figure 6.**
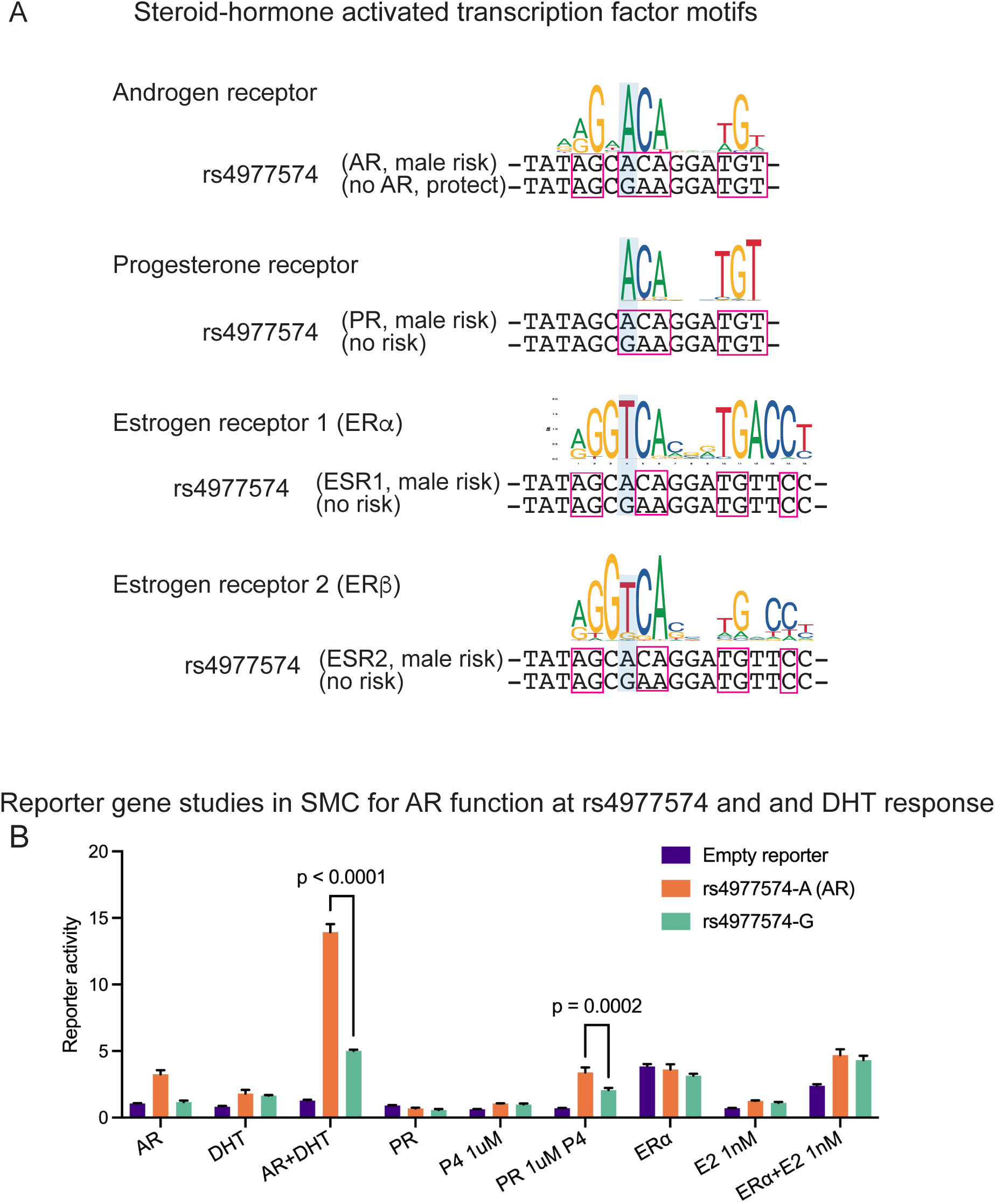
Steroid hormone activated transcription factor binding motifs. **A)** TF binding motifs and their relationship to variants at rs4977574. **B)** Transcriptional expression linked to rs4977574 with reporter gene studies in A7R5 rat SMC. DHT, dihydrotestosterone; PR, progesterone receptor; P4, progesterone; ERa, estrogen receptor alpha; E2, estradiol 1nM.

**Suppl. Figure 7.**
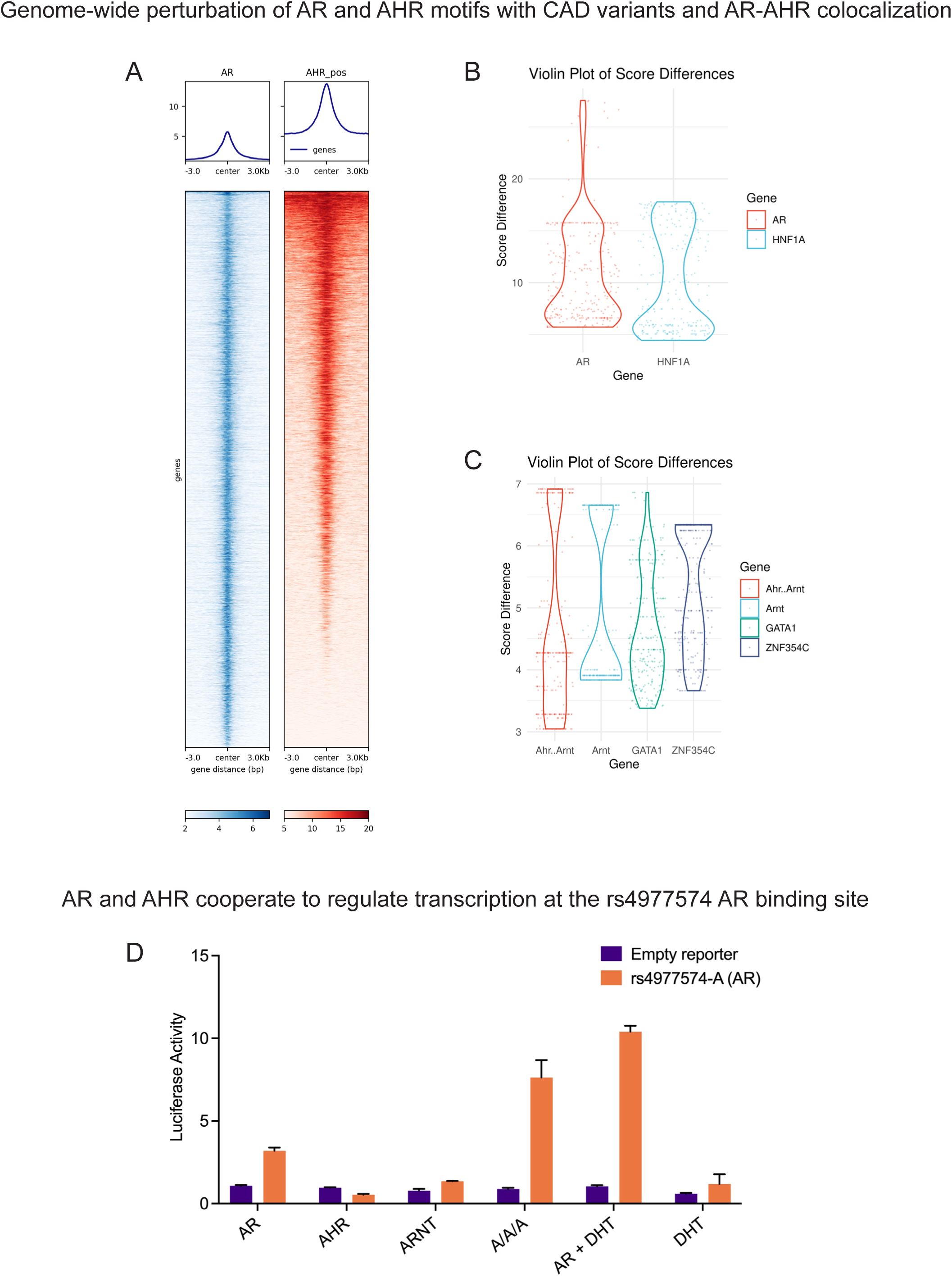
AR and AHR colocalize across the genome, and cooperate to regulate transcription. **A)** Heatmap shows AR binding regions across the human genome as determined with ChIP-seq in prostate carcinoma cells, colocalize with AHR binding sites in the same regions as identified with ChIP-seq of HCASMC (p <2.2e-16 by the Wilcoxon test). **B)** Violin plots of disruption scores show that variants in AR binding motifs are more likely to impair binding compared to other binding motifs of similar length and complexity such as shown for control TF HNF1A (p <2.2e-16) by the Wilcoxon rank sum test). **C)** Violin plots of disruption scores show that variants in AHR binding motifs are not more likely to impair binding compared to control GATA1 and ZNF354C TFs with similar length and complexity. **D)** AR and AHR cooperate to regulate transcription at AR binding sites, as shown for the rs4977574 AR binding site with luciferase reporter gene studies in A7R5 SMC. A luciferase reporter plasmid containing amplified DNA homozygous for the AR binding site at rs4977574 (A allele) was transfected into SMC with expression vectors encoding AHR, ARNT, AR, a combination of all TFs (A/A/A), AR with DHT stimulation (AR + DHT) or DHT stimulation alone with a control reporter construct.

